# Genotype-predicted drug response phenotypes and their co-occurrence with dispensed medicines among 738,531 participants in the UK Our Future Health study

**DOI:** 10.64898/2026.08.11.26360205

**Authors:** Christopher T. Rentsch, Krishnan Bhaskaran, Mirko Pavicic, Helen R. Warren, Julian Matthewman, Eleanor Barry, Imran Rafi, Judith Hayward, Clare Gerada, Anoop Shah, Patricia B. Munroe, Matt J. Silver, Munir Pirmohamed

## Abstract

Pharmacogenomics (PGx) can improve safety and effectiveness of commonly dispensed medicines, but its value at the population level depends on how often clinically actionable PGx phenotypes co-occur with the medicines they affect. We assessed this co-occurrence in a cross-sectional analysis of Our Future Health (OFH), a new UK national biobank, by applying Pharmacogenomics Clinical Annotation Tool (PharmCAT v3.1.1) to imputed genotypes from 738,531 participants across 17 pharmacogenes with established PGx prescribing guidelines. Every participant had at least one actionable PGx phenotype, with a mean of 6.1 (SD 1.3). The number of actionable PGx phenotypes was similar across genetically inferred ancestry groups, although the pharmacogenes contributing to that count differed between groups. Using linked primary care dispensing records, 36.8% (95% CI 36.7-36.9) had been dispensed at least one medicine between April 2018 and June 2025 matched to a gene for which they carried an actionable PGx phenotype. Co-occurrence rose with age, ranging from 43.7% to 58.9% across ancestry groups among those aged ≥70 years. Participants carried an actionable PGx phenotype for a mean of 13.8 (SD 6.5) of the 33 medicines dispensed in English primary care with PGx prescribing guidance, of which a mean of 0.6 (SD 1.0) had been dispensed. Co-occurrence was concentrated in a few widely dispensed classes, principally proton-pump inhibitors and antidepressants acting through *CYP2C19* and statins through *SLCO1B1*. These findings highlight opportunities to optimise treatment for a large proportion of patients receiving routine medications and identify where pre-emptive PGx testing could have the greatest clinical benefit.

**One Sentence Summary:** A third of participants had an actionable pharmacogenomic phenotype and were dispensed a gene-matched medicine, showing reach for pre-emptive testing.

## INTRODUCTION

Genetic variation in drug-metabolising enzymes, transporters, and drug targets can have a substantial impact on medication efficacy and the risk of adverse drug reactions (*1, 2*). In the UK, adverse drug reactions account for approximately 16.5% of hospital admissions at an estimated cost of £2.2 billion each year, and a subset can potentially be mitigated through pharmacogenomic (PGx)-guided prescribing (*3*). Of over 1.3 million adverse drug reactions reported to the national pharmacovigilance scheme between 1963 and 2024, 9% involved medicines for which PGx prescribing guidance could modify that risk.(*4*) International and national consortia – including the Clinical Pharmacogenetics Implementation Consortium (CPIC) (*5, 6*), the Dutch Pharmacogenetics Working Group (DPWG) (*7, 8*), and the UK Centre of Excellence in Regulatory Science and Innovation in Pharmacogenomics (CERSI-PGx) (*9*) – issue evidence-based prescribing guidance for a growing set of genes that influence drug response. Specific genotypes at these pharmacogenes can be translated into a predicted drug-response phenotype (e.g., a poor metaboliser who clears certain medicines more slowly than normal). Where such a phenotype warrants deviating from standard care (e.g., dose adjustment, avoidance of a drug, or selection of an alternative agent) it is deemed actionable; hereafter, an actionable PGx phenotype. Because many of the current guidelines affect medicines prescribed at scale for common conditions, including statins for elevated cholesterol, antidepressants for depression and anxiety, and proton-pump inhibitors for acid reflux, the potential benefits of PGx to population health are considerable.

A pre-emptive model, in which PGx information is generated before prescribing and incorporated into clinical decision-making at the point of care, reduced clinically relevant adverse drug reactions by 30% in a cluster-randomised implementation study across seven European countries (*10*). Genotype frequency alone does not establish the potential reach of a pre-emptive model; it is also necessary to know how often people carrying actionable PGx phenotypes are dispensed medicines they affect. National biobanks linking genomic data to primary care dispensing records offer an opportunity to estimate both at population scale, and thereby quantify the potential for PGx-guided prescribing. In the UK, such prescribing is beginning to move into routine care: the National Institute for Health and Care Excellence (NICE) now recommends *CYP2C19* genotyping before prescribing clopidogrel for secondary stroke prevention (*11, 12*), and National Health Service (NHS) England has begun piloting broader panel-based testing (*13*). This shifts the question to what a broader panel would yield: how many people carry actionable phenotypes, how often those phenotypes co-occur with the medicines they affect, and therefore where pre-emptive testing would have the greatest reach.

Previous large-scale studies have established that actionable PGx phenotypes are near-universal, with prevalence estimates above 98% across several major biobanks (*14–16*). Though the prevalence of actionable PGx phenotypes differs substantially between ancestry groups (*17*), current evidence is largely derived from European-ancestry populations. Far less is known about how often these genetically predicted phenotypes co-occur with actual medication exposure.

Where this has been examined, it has been limited, with incomplete ascertainment of genetic information or medication exposure, or without the individual-level information or scale to examine variation between ancestry groups. A UK Biobank study found that 24% of participants had been prescribed a medication for which they carried an actionable PGx phenotype (*14*).

However, the analysis was limited to 28,101 of its 500,000 participants with the necessary data linkages. A US study of 7.8 million Veterans Health Administration pharmacy users found that 55% had received at least one drug with a CPIC level A recommendation, though pharmacogenetic variants were projected from reference population allele frequencies weighted to the self-reported racial and ethnic composition of the cohort rather than directly assayed (*18*).

Our Future Health (OFH) is a new UK research programme established to support research into the prevention, early detection, and treatment of common diseases (*19*). Since 2021 it has recruited over two million participants, with a target of five million, and an explicit aim of recruiting a cohort that reflects the diversity of the UK population. Genome-wide imputed genotypes are currently available for 755,000 participants, and consent permits linkage to NHS records, including dispensing for medicines in primary care. To date, the only published PGx analysis in OFH examined four pharmacogenes using array data and self-reported medication use, without linkage to dispensing records(*20*). To our knowledge, no study in this cohort has combined imputed genotypes across a broader panel of guideline-backed pharmacogenes with linked dispensing data and stratification by genetically-inferred ancestry (GIA).

We therefore set out, in OFH, to (i) estimate the prevalence of actionable PGx phenotypes across pharmacogenes with established CPIC/DPWG guidance covered by the Pharmacogenomics Clinical Annotation Tool (PharmCAT), overall and by GIA; and (ii) estimate, among participants with an actionable PGx phenotype, the proportion who had been dispensed a gene-matched medicine in linked primary care dispensing records. These analyses aim to assess the potential for PGx-guided prescribing rather than its potential clinical effectiveness.

## RESULTS

### Cohort description

Imputed genotype data were available for 755,000 OFH participants. Five had no linked record in the curated participant data, 16,291 were not recruited in England, and 173 had no recorded date of birth; all were excluded, leaving 738,531 participants in the analysis cohort. Of these, 55.3% were female and median age at recruitment was 56.4 years (IQR 42.3 to 66.4) (**Table 1**). GIA was distributed as 90.6% European (EUR), 4.6% South Asian (SAS), 1.9% East Asian (EAS), 1.5% African (AFR), 1.1% Middle Eastern or North African (MENA) and 0.2% Admixed American (AMR).

**Table 1.** Cohort characteristics.

|  | **Overall** | **EUR** | **SAS** | **EAS** | **AFR** | **MENA** | **AMR** |
| --- | --- | --- | --- | --- | --- | --- | --- |
| Sample size, n | 738,531 | 669,368 | 33,935 | 13,906 | 11,379 | 8,341 | 1,602 |
| Female, n (%) | 408,511 (55.3) | 372,150 (55.6) | 16,099 (47.4) | 8,896 (64.0) | 6,042 (53.1) | 4,262 (51.1) | 1,062 (66.3) |
| Age, median (IQR) | 56.4 (42.3 to 66.4) | 57.6 (43.7 to 67.1) | 43.6 (34.4 to 55.2) | 46.0 (35.9 to 55.5) | 46.0 (35.6 to 57.5) | 43.9 (34.7 to 55.7) | 43.0 (34.3 to 51.7) |
| Age 18-39, n (%) | 159,235 (21.6) | 133,037 (19.9) | 13,529 (39.9) | 4,765 (34.3) | 3,974 (34.9) | 3,267 (39.2) | 663 (41.4) |
| Age 40-59, n (%) | 273,491 (37.0) | 242,619 (36.2) | 14,504 (42.7) | 6,807 (49.0) | 5,221 (45.9) | 3,583 (43.0) | 757 (47.3) |
| Age 60-69, n (%) | 180,999 (24.5) | 172,283 (25.7) | 4,072 (12.0) | 1,766 (12.7) | 1,732 (15.2) | 1,020 (12.2) | 126 (7.9) |
| Age 70+, n (%) | 124,806 (16.9) | 121,429 (18.1) | 1,830 (5.4) | 568 (4.1) | 452 (4.0) | 471 (5.6) | 56 (3.5) |

*Abbreviations:* EUR, European ancestry; SAS, South Asian ancestry; EAS, East Asian ancestry; AFR, African ancestry; MENA, Middle Eastern or North African ancestry; AMR, Admixed American ancestry; IQR, interquartile range
*Notes:* Age was assessed at registration into the Our Future Health study

### Prevalence of actionable PGx phenotypes

Of the 19 pharmacogenes assessed, 17 yielded analysable PGx phenotypes (16 called by PharmCAT and *CYP2C19* recovered separately; see Materials and Methods), while *CYP4F2* and *IFNL3* had no assigned PGx phenotype. At least one actionable PGx phenotype, defined as one which guidelines advise deviating from standard care, was present in all 738,531 (100%) participants. In a sensitivity analysis restricted to the 14 pharmacogenes additionally excluding those at which an actionable PGx phenotype was near-universal (i.e., *CYP3A5*, *UGT1A1*, *CFTR*), 98.9% of participants retained at least one actionable PGx phenotype. The number of actionable PGx phenotypes per participant ranged from 1 to 12, with a mean of 6.1 (standard deviation [SD] 1.3). This varied by GIA group, from 5.4 (SD 1.3) in AFR participants to 6.2 (SD 1.3) in EUR participants (**Fig. 1**)

**Figure 1.**
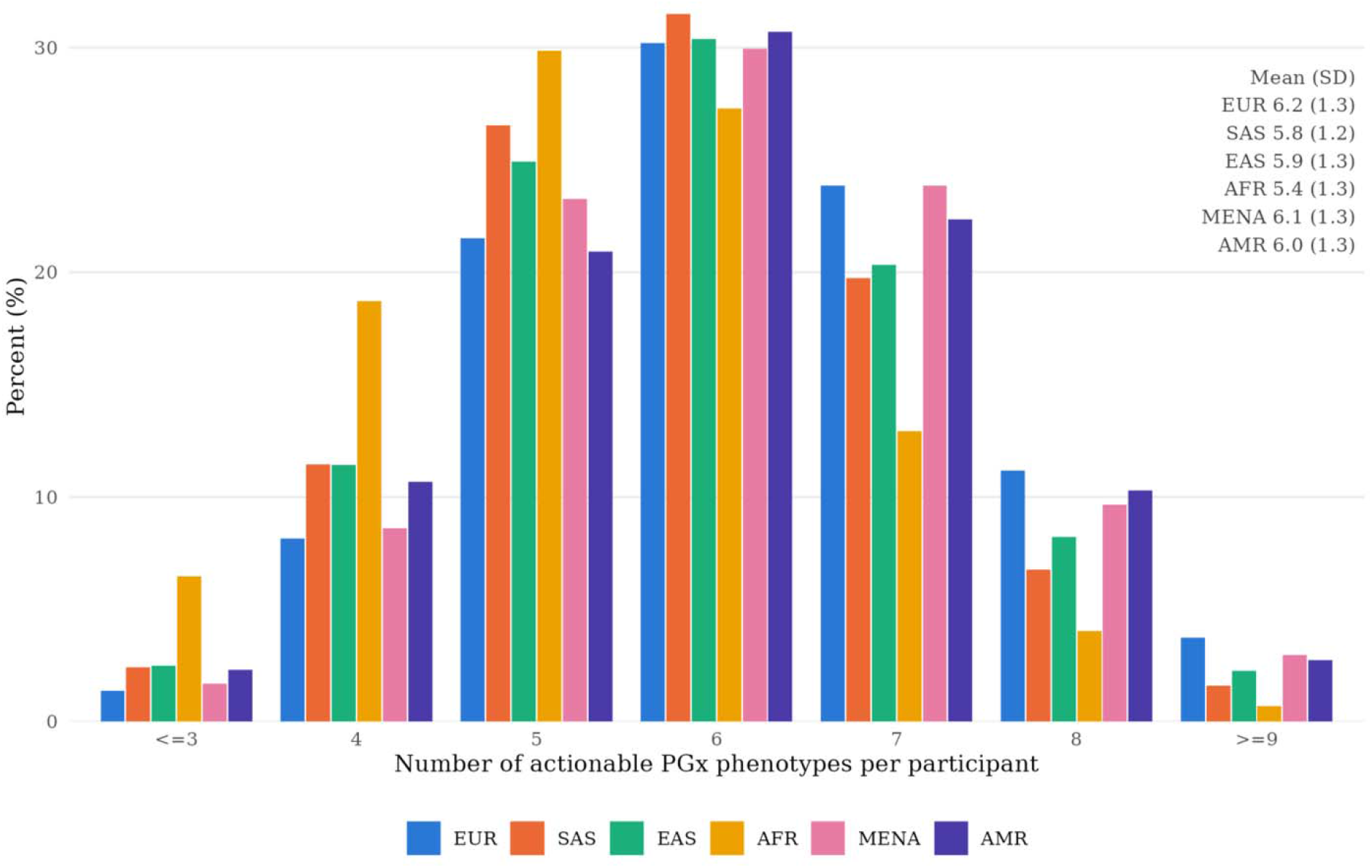
Number of actionable pharmacogenomic (PGx) phenotypes per participant by genetically-inferred ancestry group *Abbreviations:* PGx, pharmacogenomic; EUR, European ancestry; SAS, South Asian ancestry; EAS, East Asian ancestry; AFR, African ancestry; MENA, Middle Eastern or North African ancestry; AMR, Admixed American ancestry

The proportion of participants with an actionable PGx phenotype varied widely across the 17 pharmacogenes (**Table 2**). An actionable PGx phenotype was present in almost all participants for three pharmacogenes: *CYP3A5* (98.6%), *UGT1A1* (98.6%) and *CFTR* (98.1%); and in a majority for a further three: *CYP2C19* (60.7%), *VKORC1* (60.4%) and *NAT2* (59.2%).

**Table 2.** Distribution of actionability of genetically predicted drug response phenotypes across 17 pharmacogenes.

|  | Actionable | Not Actionable | Indeterminate/Missing |
| --- | --- | --- | --- |
| *ABCG2* | 153,621 (20.8) | 584,910 (79.2) | 0 (0.0) |
| *CACNA1S* | redacted | redacted | 0 (0.0) |
| *CFTR* | 724,782 (98.1) | 0 (0.0) | 13,749 (1.9) |
| *CYP2B6* | 300,816 (40.7) | 378,721 (51.3) | 58,994 (8.0) |
| *CYP2C19* | 448,285 (60.7) | 289,083 (39.1) | 1,163 (0.2) |
| *CYP2C9* | 163,447 (22.1) | 447,061 (60.5) | 128,023 (17.3) |
| *CYP3A4* | 60,881 (8.2) | 659,087 (89.2) | 18,563 (2.5) |
| *CYP3A5* | 728,021 (98.6) | redacted | redacted |
| *DPYD* | 46,655 (6.3) | 691,876 (93.7) | 0 (0.0) |
| *G6PD* | 4,217 (0.6) | 728,586 (98.7) | 5,728 (0.8) |
| *NAT2* | 437,408 (59.2) | 0 (0.0) | 301,123 (40.8) |
| *NUDT15* | 13,081 (1.8) | 718,800 (97.3) | 6,650 (0.9) |
| *RYR1* | 127 (0.0) | 738,404 (>99.9) | 0 (0.0) |
| *SLCO1B1* | 200,248 (27.1) | 529,731 (71.7) | 8,552 (1.2) |
| *TPMT* | 68,831 (9.3) | 667,335 (90.4) | 2,365 (0.3) |
| *UGT1A1* | 728,312 (98.6) | 3,386 (0.5) | 6,833 (0.9) |
| *VKORC1* | 445,781 (60.4) | 292,750 (39.6) | 0 (0.0) |

*Notes:* Genetically predicted drug response phenotypes were assigned by Pharmacogenomics Clinical Annotation Tool (PharmCAT v3.1.1) and classified as actionable when guidelines would advise deviating from standard care. Percentages are within gene. Cells are marked redacted to protect small cell counts; more than one cell may be redacted within a gene for this purpose, so a redacted cell does not always indicate a small cell count. Two genes (*CYP4F2* and *IFNL3*) had no assigned phenotype and were excluded from all analyses. PharmCAT assigns no “Normal Metabolizer” phenotype for *NAT2*; all participants with a determinate call are therefore actionable.

Intermediate proportions were seen for *CYP2B6* (40.7%), *SLCO1B1* (27.1%), *CYP2C9* (22.1%) and *ABCG2* (20.8%), and lower proportions for *TPMT* (9.3%), *CYP3A4* (8.2%), *DPYD* (6.3%), *NUDT15* (1.8%) and *G6PD* (0.6%). An actionable PGx phenotype was rare for *CACNA1S* and *RYR1* (<0.1%). The specific genetically predicted drug response phenotypes underlying each actionable call for each pharmacogene are given in **Table S1**. For *CYP2C19*, for example, the 60.7% actionable total spans poor (3.0%), intermediate (27.3%), rapid (25.9%), and ultrarapid (4.5%) metaboliser phenotypes, reflecting both reduced-and increased-function genotypes.

The proportion of participants with an actionable PGx phenotype differed substantially between GIA groups, and the direction and magnitude of these differences were gene-specific. For some pharmacogenes, the actionable proportion was high across all groups: *CFTR* exceeded 97% in every group (**Fig. 2**, **Table S2**). For others it varied several-fold. An actionable *CYP3A5* phenotype was present in 99.5% of EUR participants and 68.2% of AFR participants; conversely, actionable *VKORC1* phenotypes were most common in EAS (97.8%) and least common in AFR (15.1%). *ABCG2* ranged from 2.8% (AFR) to 49.1% (EAS), and *G6PD*, though uncommon overall, was most frequent in AFR (9.7%). The full distribution of genetically predicted drug response phenotypes by GIA group are presented in **Table S3**.

**Figure 2.**
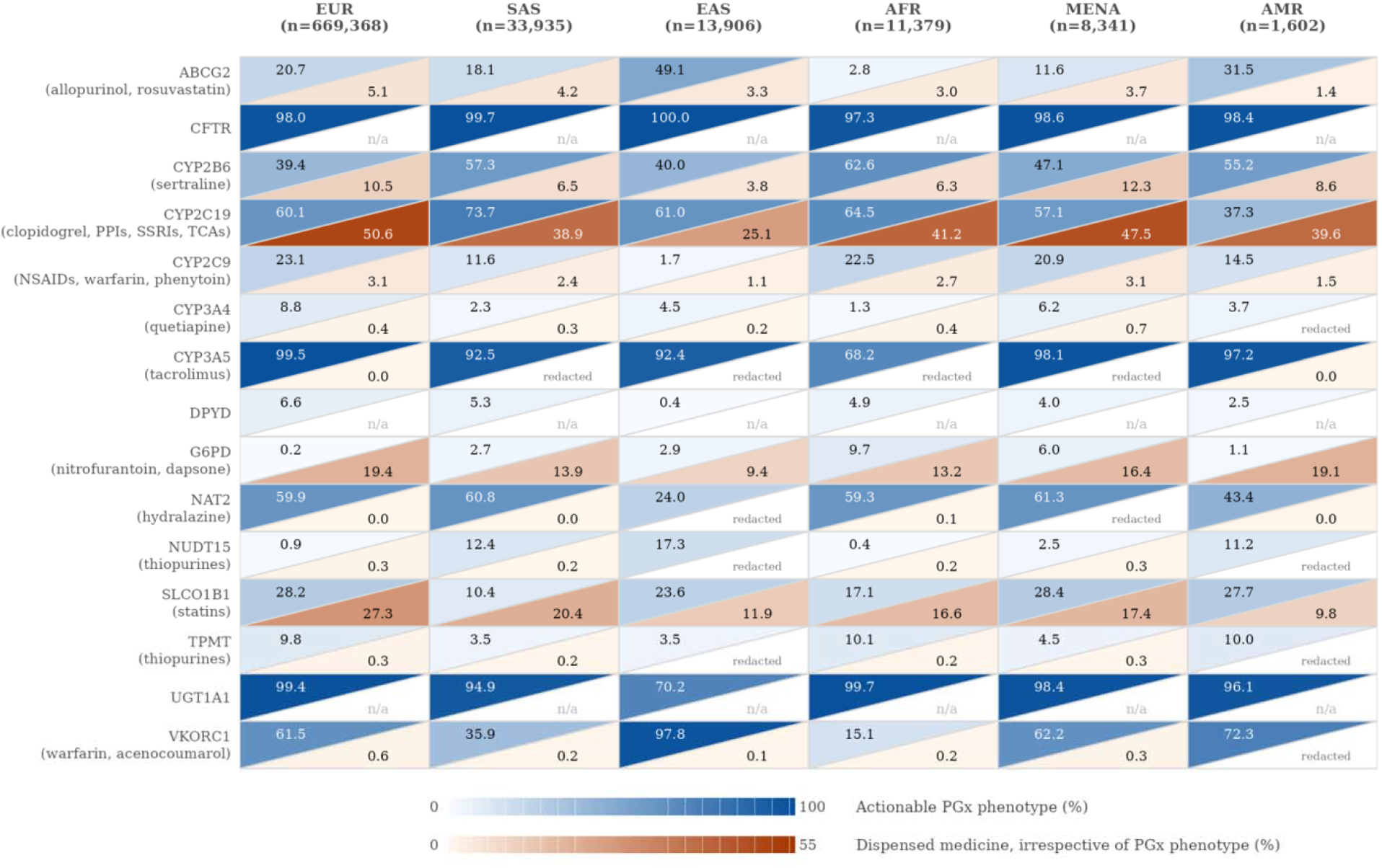
Prevalence of actionable pharmacogenomic (PGx) phenotypes and dispensing of paired medicines, by gene and genetically-inferred ancestry group *Abbreviations:* PGx, pharmacogenomic; EUR, European ancestry; SAS, South Asian ancestry; EAS, East Asian ancestry; AFR, African ancestry; MENA, Middle Eastern or North African ancestry; AMR, Admixed American ancestry; n/a, not applicable due to the gene having no paired medicine prescribed in primary care; PPIs, proton pump inhibitors; SSRIs; selective serotonin reuptake inhibitors; TCAs, tricyclic antidepressants; NSAID, non-steroidal anti-inflammatory drugs *Notes:* Each cell is divided diagonally: the upper left (blue scale) gives the percentage of the ancestry group with an actionable PGx phenotype at that gene, the lower right (orange scale) gives the percentage of the ancestry group dispensed at least one medicine paired with that gene, irrespective of the participants’ PGx phenotype. The two are computed over the same denominator but are not subsets of one another. Dispensing is drawn from linked NHS Business Services Authority primary care records covering April 2018 to June 2025. A participant dispensed more than one medicine paired with the same gene is counted once. A participant can be counted across multiple genes. The set of medicines include 39 gene-drug pairs comprising 33 distinct medicines dispensed in primary care. Of the 19 genes covered in PharmCAT v3.1.1, two genes *(CYP4F2* and *IFNL3)* had no assigned phenotype and were excluded from all analyses. Two additional genes *(CACNAIS* and *RYRI)* were excluded due to small cell sizes in all ancestry groups. Three genes *(CFTR, DPYD,* and *UGT1A1)* have no paired medicine dispensed in primary care and are marked “n/a”. Cells are marked “redacted” to protect small cell counts; more than one cel! may be redacted within a gene for this purpose, so a redacted cell does not always indicate a small cell count.

### Dispensing of medicines with actionable prescribing guidance

Overall, 469,824 (63.6%, 95% CI 63.5 to 63.7) participants had been dispensed at least one medicine carrying an actionable gene-drug recommendation between April 2018 and June 2025, irrespective of their PGx phenotype at the paired gene. Dispensing was most common in EUR participants (65.1%) and least common in EAS (36.7%), with intermediate proportions in MENA (57.2%), AFR (51.2%), SAS (51.0%) and AMR (50.9%). Dispensing was highest for medicines paired with *CYP2C19*, including proton-pump inhibitors, some antidepressants, and clopidogrel, ranging from 25.1% to 50.6% across GIA groups (**Fig. 2**, **Table S4**). Statins acting through *SLCO1B1* ranged from 9.8% to 27.3%, nitrofurantoin through *G6PD* from 9.4% to 19.4%, and sertraline through *CYP2B6* from 3.8% to 12.3%. For all other pharmacogenes with a medicine dispensed in primary care, fewer than 5.1% of participants in any GIA group were dispensed a paired medicine.

### Co-occurrence of actionable PGx phenotypes with dispensed medicines

Overall, 271,510 (36.8%, 95% CI 36.7 to 36.9) participants had been dispensed at least one medicine between April 2018 and June 2025 matched to a gene for which they carried an actionable PGx phenotype. Co-occurrence varied by GIA group, from 37.6% (95% CI 37.5 to 37.7) in EUR participants to 19.0% (18.3 to 19.6) in EAS (**Fig. 3**). Within every GIA group, co-occurrence at least doubled between participants aged 18 to 39 years and those aged 70 years or above, from a range of 12.0% to 26.6% in the youngest band to 43.7% to 58.9% in the oldest (**Fig. 3**).

**Figure 3.**
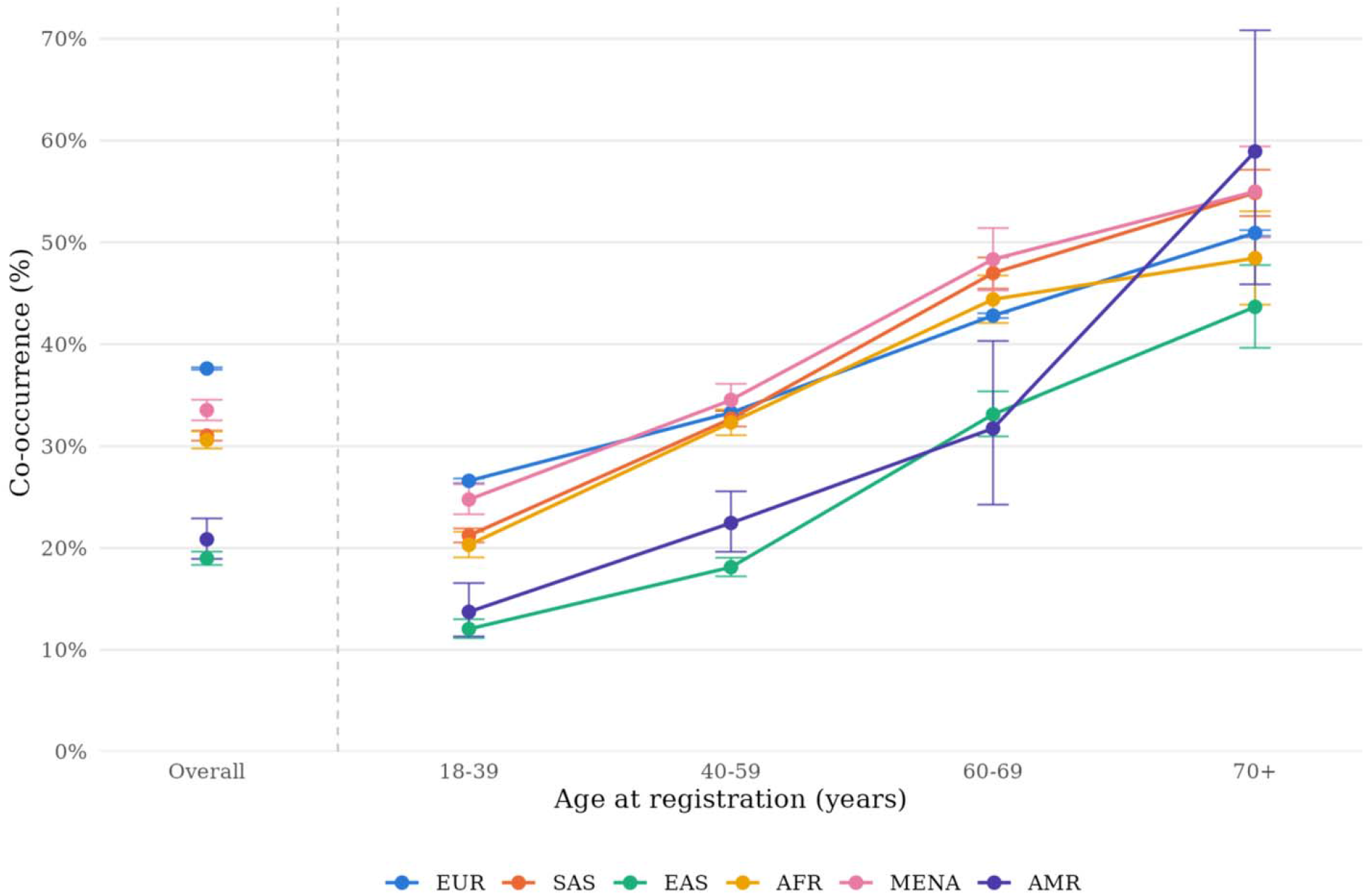
Co-occurrence of actionable pharmacogenomic (PGx) phenotypes and gene-matched dispensed medicines, by age at registration and genetically-inferred ancestry group *Abbreviations:* PGx, pharmacogenomic; EUR, European ancestry; SAS, South Asian ancestry; EAS, East Asian ancestry; AFR. African ancestry; MENA, Middle Eastern or North African ancestiy; AMR, Admixed American ancestry *Notes:* Dispensing is drawn from linked NHS Business Services Authority primary care records covering April 2018 to June 2025. A participant dispensed more than one medicine paired with the same gene is counted once. A participant can be counted across multiple genes. The set of medicines include 39 gene-drug pairs comprising 33 distinct medicines dispensed in primary care. Three genes *(CFTR, DP YD.* and *UGT1A1)* have no paired medicine prescribed in primary care. 95% confidence intervals were calculated using the Wilson score method without continuity correction.

Across the 80 gene-drug pairs with an actionable recommendation, comprising 66 distinct medicines, participants carried an actionable PGx phenotype for a mean of 22.4 medicines (SD 8.7; range 1 to 55). Restricting to the 39 gene-drug pairs comprising 33 distinct medicines dispensed in English primary care, the mean was 13.8 (SD 6.5; range 0 to 31). Of these, participants were actually dispensed a mean of 0.6 (SD 1.0; range 0 to 10).

Co-occurrence was largely driven by a few widely prescribed drug classes. Of the 448,285 participants with an actionable *CYP2C19* phenotype, 120,809 (26.9%) were dispensed omeprazole (**Fig. 4**). The proportion was also high for *SLCO1B1*-atorvastatin (23.2%), *CYP2C19*-lansoprazole (18.4%), *G6PD*-nitrofurantoin (16.7%), *CYP2C19*-amitriptyline (12.1%), *CYP2C19*-sertraline (10.1%), and *CYP2B6*-sertraline (10.0%). Seven more gene-drug pairs were observed with a percentage between 1.6% and 5.9%, and the remainder were below 1.0%.

**Figure 4.**
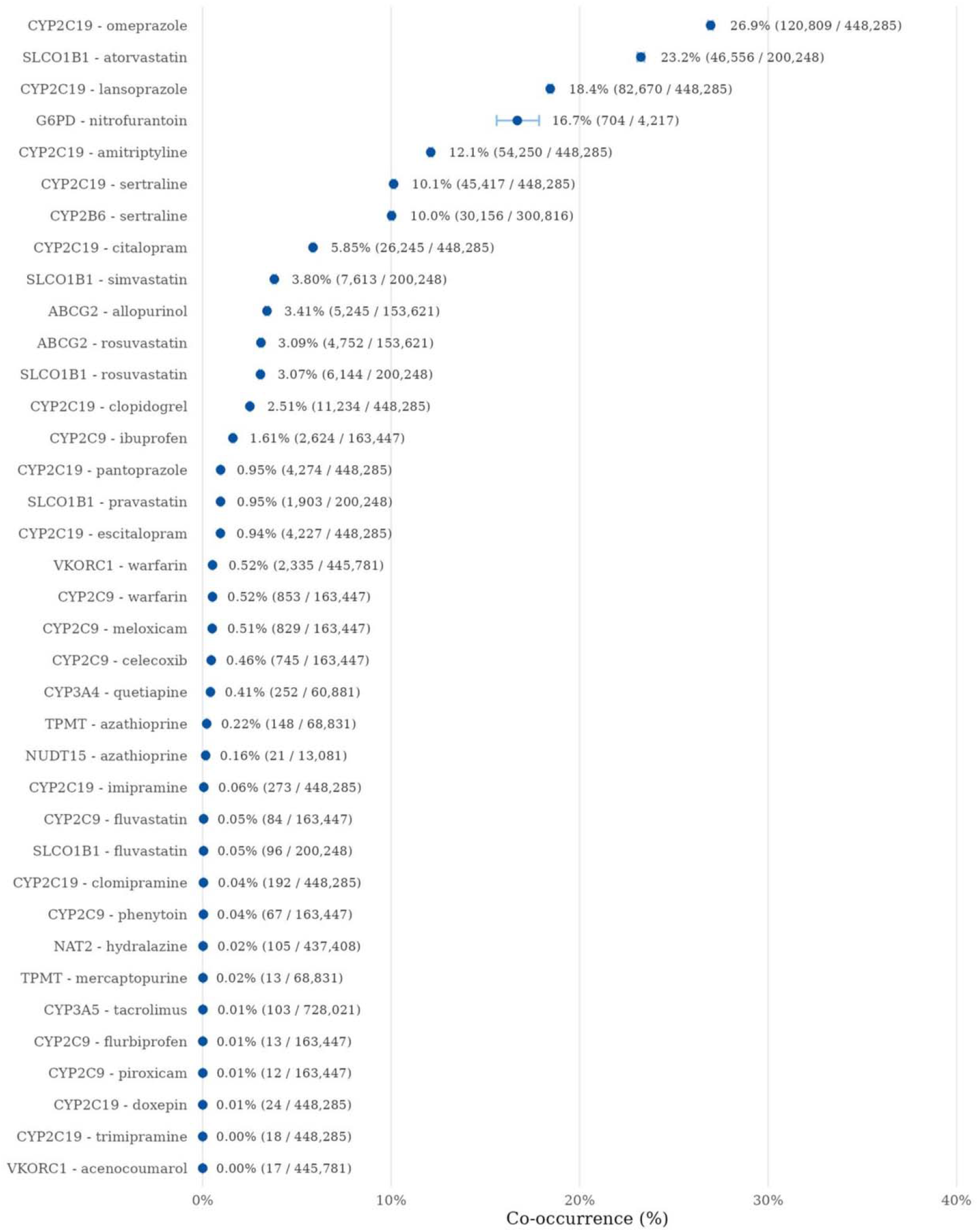
Co-occurrence of an actionable pharmacogenomic (PGx) phenotype and receipt of a gene matched medicine, by gene-drug pair *Abbreviations:* PGx, pharmacogenomic *Notes:* For each gene-drug pair, the denominator is participants with an actionable PGx phenotype at the gene; the numerator is those among them with a dispensing record for the paired medicine. Dispensing is drawn from linked NHS Business Services Authority primary care records covering April 2018 to June 2025. A participant can be counted across multiple gene-drug pairs. This figure covers 39 gene-drug pairs comprising 33 distinct medicines dispensed in primary care. Of these, two are not displayed: GWVJ-dapsone and *NUDT15-*mercaptopurine are suppressed due to counts of participants dispensed the medicines lower than the disclosure threshold (<l 0). 95% confidence intervals were calculated using the Wilson score method without continuity correction.

## DISCUSSION

In this analysis of 738,531 participants in a new UK national biobank, more than a third had been dispensed a medicine matched to a gene for which they carried an actionable PGx phenotype.

That co-occurrence was concentrated in a small number of high-volume primary care medicines, principally proton-pump inhibitors and antidepressants acting through *CYP2C19* and statins through *SLCO1B1*. Actionable PGx phenotypes were universal: every participant carried at least one, with a mean of 6.1, consistent with previous biobank studies. The scale of OFH and its linkage to dispensing records enabled the discovery of where the co-occurrence of actionable PGx phenotypes and gene-matched primary care dispensing is currently greatest, and therefore which gene-drug pairs future safety, effectiveness, and implementation studies could prioritise.

A distinct contribution of this study was the quantification of co-occurrence between actionable PGx phenotypes and the medicines they affect across a large set of pharmacogenes with established PGx prescribing guidelines. We found that more than 1 in 3 OFH participants had both an actionable PGx phenotype and a dispensing record for the corresponding medicine, equivalent to a mean of 0.6 medicines per participant. Related estimates have been reported in other biobanks: approximately 1 in 4 in UK Biobank (*14*), 1 in 5 in All of Us (*15*), and 1 in 7 in the PennMedicine Biobank (*16*). Even lower proportions have been reported in smaller cohorts assessing a narrower set of gene-drug pairs (*21*). However, these figures are not directly comparable. They differ in how completely medication exposure was ascertained, and in the prescribing patterns and age structures of the populations studied. Co-occurrence is therefore present across independent healthcare systems, and its measured magnitude reflects the population and the data available rather than underlying genetic variation.

Our finding that all participants carried at least one actionable PGx phenotype is consistent with prior biobank analyses, whether based on imputed genotypes, genotyping arrays, exome, or whole-genome sequence data: 99.5% in the UK Biobank (*14*), 100% in the All of Us Research Program (*15*), and 98.9% in the Penn Medicine BioBank (*16*). The same held when expressed at the level of individual medicines: OFH participants carried an actionable PGx phenotype for a mean of 13.8 of the 33 medicines dispensed in English primary care with established PGx prescribing guidelines, broadly comparable with the mean of 10.3 medicines with a predicted atypical response reported in the UK Biobank given the latter analysis included a smaller set of genes (*14*). That we produced both of these from imputed data, and recovered phenotype calls for a major pharmacogene, *CYP2C19*, that would otherwise have been lost to a panel gap, supports the use of imputation-based approaches for population-scale PGx characterisation.

Across all these large-scale biobanks, actionability at the “any gene” level is saturated.(*22*) In our study, this saturation did not depend on genes at which actionable PGx phenotypes were near-universal. The more discriminating question for implementation is therefore not whether a participant carries an actionable PGx phenotype, but how many they carry, and for which medicines.

Co-occurrence varied by GIA group, from 37.6% in EUR to 19.0% in EAS participants. Reporting dispensing separately from PGx phenotype allowed their contributions to co-occurrence to be partly distinguished. While the mean number of actionable PGx phenotypes varied little across GIA groups (5.4 to 6.2), dispensing varied nearly two-fold (36.7% to 65.1%). Variation in co-occurrence therefore tracked variation in dispensing more than in the prevalence of actionable PGx phenotypes, and dispensing is largely shaped by prescribing patterns, disease prevalence, healthcare access, and the younger age structure of non-EUR groups in OFH.

When stratified by age, co-occurrence rose steeply across all GIA groups, at least doubling from the youngest to the oldest bands. Two explanations cannot be disentangled in these data. Older participants are more likely to be prescribed these medicines, reflecting greater comorbidity and polypharmacy. They have also had more opportunity to be dispensed any medicine within a fixed seven-year window. Both factors support our estimates being a lower bound on lifetime co-occurrence, particularly for older participants, for whom the window captures only a short segment of a much longer prescribing history.

Although the overall number of actionable PGx phenotypes was similar across GIA groups, the specific pharmacogenes contributing to that count differed between groups. These differences require careful interpretation. Lower actionability for particular genes in particular groups (e.g., *CYP3A5* and *VKORC1* in participants of African ancestry) likely reflects the composition of the current guidelines and the EUR-skewed evidence base from which it derives, not a lesser need for PGx-guided prescribing in those participants. Consistent with this, non-EUR populations have been shown to carry a greater frequency of functionally consequential variants that are not captured by current star-allele definitions (*14, 17, 23*), meaning that our estimates, in common with those of other biobank studies, may understate actionability in non-EUR groups. As PGx testing moves toward implementation, reference resources and allele definitions that perform equitably across ancestries will be essential if it is to narrow rather than widen existing inequities in the evidence base.

This study has several strengths. First, it was conducted in a large-scale biobank cohort recruited with an explicit commitment to diversity. Although participants of EUR ancestry remained the large majority, the scale of the cohort meant that the non-EUR groups were large enough to yield stable estimates, including for groups often too small for stratified analysis in other datasets. As genotyping extends to the full recruited cohort, these groups will grow substantially, supporting more granular ancestry-specific analysis than is currently possible in any comparable biobank.

Second, ancestry was assigned using a population-scale genetic method rather than relying on self-reported ethnicity. Third, we characterised phenotypes across all CPIC/DPWG guideline-backed genes covered by PharmCAT v3.1.1, and linked them to dispensing records rather than basing exposure on self-reported use.

The study also has limitations. First, imputed genotypes could not resolve all star alleles; rare alleles that depend on positions absent from the imputation panel remained uncallable, and *CYP2C19* required a bespoke recovery procedure. Second, several clinically important pharmacogenes could not be assessed. For *CYP4F2* and *IFNL3*, PharmCAT v3.1.1 assigned no metaboliser phenotype, so no actionability could be determined despite genotypes and diplotypes being called successfully; *CYP4F2*3* has been associated with higher warfarin maintenance dose requirements,(*24*) and any actionability at these genes is therefore unaccounted for in our estimates. *CYP2D6*, which contributes to the metabolism of several high-volume medicines, including codeine, tamoxifen, and many antidepressants, is not reliably resolved because of its structural and copy-number variation.(*25*) PharmCAT does not call *CYP2D6* from imputed data, requiring specialised callers and sequence data.(*26*) The same constraint applies to *HLA* loci that carry some of the strongest pharmacogenetic associations in clinical use (e.g., *HLA-A* for carbamazepine and *HLA-B* for abacavir).(*27*) A further pharmacogene, *ACKR1*, which influences baseline neutrophil counts and is of particular importance for AFR and MENA populations,(*28*) is not currently covered by PharmCAT. Because these genes include several highly actionable associations – *CYP2D6* among the three pharmacogenes accounting for most pharmacogenomically modifiable adverse drug reactions reported in the UK (*4*) – their omission means that we likely have understated the potential reach of PGx-guided prescribing. Third, the dispensed medications data in OFH captures community dispensing between April 2018 and June 2025 in England only; thus, OFH participants recruited in Scotland, Wales and Northern Ireland were excluded. Private prescriptions (unusual given the NHS), medicines only administered in secondary care, and over-the-counter purchases such as ibuprofen were not captured, so exposure was incompletely ascertained for some medicines. Country of recruitment was used as a proxy for the English dispensing catchment, and some misclassification is possible. Importantly, this dispensing window spans only seven years within a cohort recruited across a wide age range, so for older participants in particular it captures a short segment of a much longer prescribing history; medicines dispensed before the window were unobserved. Our co-occurrence estimates should therefore be read as a lower bound. As OFH matures, linkage to primary care records – to which participants consent at recruitment – will extend both the period and completeness of medication history. Although these records capture prescribing rather than dispensing, they will support a fuller assessment of how often actionable PGx phenotypes and gene-matched medicines co-occur. Fourth, OFH participants are not a representative sample of the UK population, therefore estimates may not be generalisable. This applies particularly to the co-occurrence analyses, which depend on both who volunteers and how they engage with primary care. Fifth, ancestry was summarised using continental groupings, which are a coarse representation of genetic variation. Treating ancestry as a continuous measure is likely a more appropriate approach as methods and cohort size allow. Finally, the co-occurrence we describe was cross-sectional. Our analysis established that actionable PGx phenotypes and gene-matched dispensing co-occur, but not that PGx-guided prescribing would have necessarily improved outcomes. Actionability was defined against current guidance, which will continue to evolve.

In conclusion, actionable PGx phenotypes were universal in this new national cohort, and more than a third of participants had been dispensed a medicine whose response they influence, despite only a seven-year dispensing window. Authoritative bodies, including the Royal College of Physicians, the British Pharmacological Society, and the European Medicines Agency, have called for the use of population-scale biobanks to strengthen the PGx evidence base (*29, 30*).

This analysis contributes to that evidence base and identifies where the co-occurrence of actionable PGx phenotypes and primary care dispensing is currently greatest, which is where future studies of safety and effectiveness could focus. What remains unresolved is whether acting on this information improves outcomes, and for whom. Continued linkage of this cohort to primary care records will support answering those questions.

## MATERIALS AND METHODS

### Study design

This was a cross-sectional assessment within a new national biobank, with two objectives: (i) to estimate the prevalence of actionable PGx phenotypes across the guideline-backed pharmacogenes covered by PharmCAT, overall and by GIA group; and (ii) among participants with an actionable PGx phenotype, to estimate the proportion dispensed the gene-matched medicine in linked primary care dispensing records. This study is reported in accordance with the Strengthening the Reporting of Observational Studies in Epidemiology (STROBE) guideline for cross-sectional studies.

### Study population and data sources

Our Future Health (OFH) is a UK research programme that has recruited over two million adult participants since 2021 (target five million), with informed consent for genome-wide genotyping and for linkage to NHS health records (*19*). For this study, we included all participants with imputed genetic data and excluded those with missing participant data or no recorded date of birth. Because linked primary care data is currently restricted to England, we only included participants recruited in England. Ethics approval for Our Future Health was granted by the Cambridge East Research Ethics Committee (#21/EE/0016). Analyses for the present study were performed within the OFH Trusted Research Environment under study ID OFHS240171 with ethical approval from London School of Hygiene & Tropical Medicine (#32534). Participants were genotyped genome-wide and imputed centrally by OFH (*31*); we used the v6 release (GRCh38, May 2026).

### Pharmacogenomic phenotyping

Pharmacogenomic phenotypes were assigned using PharmCAT v3.1.1, as previously described (*32–34*). Briefly, imputed genotypes at the variant positions defining each pharmacogene’s star alleles were used to assign each participant’s two haplotypes. The resulting pair of star alleles (i.e., diplotype) was translated into a predicted phenotype using definitions bundled with PharmCAT (e.g., a *CYP2C19* *2/*2 diplotype yields a poor metaboliser phenotype). We included all 19 pharmacogenes covered by PharmCAT v3.1.1 (*ABCG2*, *CACNA1S*, *CFTR*, *CYP2B6*, *CYP2C19*, *CYP2C9*, *CYP3A4*, *CYP3A5*, *CYP4F2*, *DPYD*, *G6PD*, *IFNL3*, *NAT2*, *NUDT15*, *RYR1*, *SLCO1B1*, *TPMT*, *UGT1A1*, *VKORC1*). PharmCAT was run in its reference container within the OFH Trusted Research Environment, processing participants in batches for tractability at scale. Of the 19 pharmacogenes assessed, PharmCAT returned a phenotype call for 16 through the standard pipeline. Two genes yielded no phenotype call: *IFNL3* and *CYP4F2*, for which genotypes and diplotypes were called but PharmCAT assigned no metaboliser phenotype. The guideline drug for *CYP4F2* is warfarin, which is also covered here through *VKORC1* and *CYP2C9*, and *IFNL3* has no CPIC or DPWG prescribing guideline. A further gene, *CYP2C19*, returned a universal no-call from the standard pipeline but was recovered under additional assumptions described below. Of the 17 pharmacogenes with phenotype calls derived from the standard pipeline or recovery procedure, per-gene rates of indeterminate or missing phenotypes were highest for *NAT2* (40.8%) and *CYP2C9* (17.3%); all others were below 8.0% (**Table S1**).

### *CYP2C19* recovery procedure

PharmCAT returned a universal no-call for *CYP2C19* because 9 of the 35 positions used to define *CYP2C19* star alleles were absent from the imputed data; their absence was confirmed across all chromosome 10 imputation batches. The 9 missing positions are required only by rare alleles (*7, *14, *18, *19, *25, *26, *29, *31, *32) and by the reference (*1) allele, whereas the core positions defining the common, clinically important alleles (including *2, *3 and *17) were observed. We therefore recovered *CYP2C19* by extracting per-sample genotypes at the *CYP2C19* positions from the imputed variant call files and matching each haplotype against the set of clinically active alleles whose core positions were fully observed, assuming the reference base at the 9 missing positions. Normal-function alleles were excluded from matching because misassigning them to *1 has no clinical consequence, both yielding a normal metaboliser phenotype; uncertain function alleles were likewise excluded as non-actionable. Where no active allele matched, *1 was assigned by exclusion. For some participants, the available positions could not distinguish *35 from *2, yielding an ambiguous call listing both. Because *35 is a no-function allele, treated identically to *2 for phenotype assignment, these were resolved by removing *35 and re-deriving the diplotype and predicted phenotype from the remaining alleles. We then reported a single diplotype where one remained and classed the call as indeterminate where ambiguity persisted. Of note, the *35 allele is near-absent in European and Asian populations and only as high as 3% in African populations (*12*). This recovery procedure yielded a *CYP2C19* phenotype for 737,368 (99.8%) participants. Within each GIA group, the recovered phenotype distributions were consistent with published allele-and phenotype-frequency data for the corresponding populations (*12*).

### Definition of actionability

A PGx phenotype was classed as actionable where current prescribing guidelines advise deviating from standard care for at least one medicine; for example, a dose adjustment, avoidance of the drug, or selection of an alternative agent. The specific genetically predicted drug response phenotypes underlying each actionable call are given in **Table S1**. Briefly, all phenotypes other than “Normal” or “Indeterminate/Missing” were classified as actionable, with the following exceptions. *VKORC1*-1639 GG was treated as not actionable, while *VKORC1* - 1639 AA and AG were actionable. *SLCO1B1* increased function was not actionable per current CPIC guidelines (*35*). *CACNA1S* and *RYR1* “Uncertain Susceptibility” were treated as not actionable, whereas “Malignant Hyperthermia Susceptibility” was actionable. For *CFTR*, both the modulator-responsive and non-responsive phenotypes were treated as actionable, because each alters prescribing decisions. Conservative mappings were applied for *CYP2C19* (ambiguous “Likely” outputs classed as indeterminate) and for *NUDT15* and *TPMT* (“Possible” classed as indeterminate). PharmCAT assigns no “Normal” phenotype for *NAT2*; all participants with a determinate call are therefore actionable. Compound phenotype strings were assigned a single label where all tokens agreed and classed as indeterminate where mixed.

### Genetically-inferred ancestry

We used GIA groupings provided centrally to all researchers by OFH, which are described in detail in the documentation (*36*). Briefly, GIA was inferred based on principal components analysis, which returned continuous admixture proportions across 25 sub-continental regions and a single hard-called ancestry label per participant. For the primary analysis we mapped the 25 hard-called regions to six continental groups (i.e., EUR, SAS, EAS, AFR, MENA and AMR) as detailed in **Table S5**, with North African assigned to MENA. Following the National Academies of Sciences, Engineering, and Medicine recommendations (*23*), continental groupings are used pragmatically to examine heterogeneity in PGx phenotype prevalence and are not interpreted as biological categories.

### Gene-drug pairs and dispensed-medication linkage

Candidate gene-drug pairs were obtained from Clinical Pharmacogenomics Resource (ClinPGx) clinical resource (*37*), which integrates guidelines issued from CPIC and the DPWG. Of 200 candidate pairs across all 19 genes captured in PharmCAT v3.1.1, we retained 81 for which CPIC or DPWG issues a prescribing recommendation. One pair (*CYP4F2*-warfarin) was excluded because PharmCAT v3.1.1 did not assign a phenotype for *CYP4F2*, leaving 80 pairs across 17 pharmacogenes. Dispensing was ascertained from the NHS Business Services Authority (NHSBSA) Medicines Dispensed in Primary Care dataset, which records medicines dispensed between 1 April 2018 and 1 June 2025 in the community in England and excludes private prescriptions and medicines administered in secondary care and not continued in primary care. Each medicine was mapped to its British National Formulary (BNF) chemical-substance code, and dispensing was identified by prefix-matching the 9-character chemical-substance code to presentation-level codes in the dataset (**Table S6**). Of the 80 gene-drug pairs with actionable guidelines, 39 corresponded to medicines dispensed in English primary care and formed the primary analysis set, comprising 33 distinct medicines paired with 12 of the 17 pharmacogenes. Of the remaining 41, 14 were rarely dispensed (fewer than 1000 prescriptions across all primary care practices in England between June 2021 and May 2026, according to OpenPrescribing) (*38*), and 27 were predominantly theatre-administered anaesthetic agents, hospital-administered oncology or specialist medicines, and medicines not routinely dispensed in English primary care. For each retained gene-drug pair, the denominator was participants with an actionable PGx phenotype for the relevant gene-drug pair and the numerator those with at least one dispensing of the paired medicine. The full list of gene-drug pairs is provided in **Table S6**.

## Statistical analysis

The prevalence of PGx phenotypes and actionability were summarised as counts and percentages with two-sided 95% confidence intervals calculated using the Wilson score method without continuity correction, overall and stratified by GIA group. No formal hypothesis tests of between-group differences were pre-specified; comparisons by GIA group are descriptive.

We derived two per-participant counts. First, the number of actionable PGx phenotypes is the number of pharmacogenes at which a participant carried an actionable PGx phenotype, out of the 17 assessed. Second, the number of medicines affected by an actionable PGx phenotype is the number of distinct medicines for which a participant carried an actionable PGx phenotype at the paired pharmacogene, deduplicating medicines paired with more than one pharmacogene so that each medicine was counted once; this was computed across all 80 gene-drug pairs with an actionable recommendation, comprising 66 distinct medicines, and again across the 39 pairs dispensed in English primary care, comprising 33 distinct medicines. Both counts were summarised as the mean (SD).

Dispensing was summarised in two ways. First, irrespective of PGx phenotype, we calculated the proportion of participants dispensed at least one medicine carrying an actionable gene-drug recommendation, overall, by GIA group, and by pharmacogene. Second, co-occurrence was defined as a participant having both an actionable PGx phenotype at a pharmacogene and a dispensing record for a medicine paired with that same pharmacogene. Co-occurrence was calculated overall, by GIA group, by gene-drug pair, and within each GIA group, by age at registration, in bands of 18 to 39, 40 to 59, 60 to 69, and 70 years or above. Age at registration was calculated as the interval between date of birth and date of registration. We additionally counted, per participant, the number of primary care medicines that were both affected by an actionable PGx phenotype and actually dispensed to that participant.

As a sensitivity analysis, we repeated the estimate of the proportion of participants with at least one actionable PGx phenotype across a restricted set of 14 pharmacogenes, excluding the two for which no phenotype could be assigned (*CYP4F2*, *IFNL3*) and the three at which an actionable phenotype was present in almost all participants (*CFTR*, *CYP3A5*, *UGT1A1*).

Analyses used R version 4.4.2, with code available at https://github.com/ehr-lshtm/ofh-pharmcat-755k. In accordance with the disclosure control requirements of the OFH Trusted Research Environment, cells including fewer than 10 participants are redacted. Suppression was applied jointly across rows and/or columns so that redacted values cannot be recovered by subtraction; a redacted cell therefore does not necessarily denote a small count.

## Data Availability

Our Future Health data cannot be shared by the authors but are available to approved researchers via the Our Future Health Trusted Research Environment; access conditions are described at https://research.ourfuturehealth.org.uk/apply-to-access-the-data/.

## Acknowledgments

This study makes use of de-identified data held by Our Future Health. We would like to acknowledge all the research participants who have donated their data to the Our Future Health research programme. The responsibility for the interpretation of the information supplied by Our Future Health is the authors’ alone. This work uses data that has been provided by patients and collected by the NHS as part of their care and support. The data are collated, maintained and quality assured by the National Disease Registration Service, which is part of NHS England. Access to the data was facilitated by the NHS England Data Access Request Service.

## Author contributions

Conceptualization: CTR, MP

Methodology: CTR, KB, MJS, MP

Investigation: CTR

Visualization: CTR

Funding acquisition: CTR, MP

Project administration: CTR, JM, EB

Supervision: CTR, KB, MP

Writing – original draft: CTR, MP

Writing – review & editing: CTR, KB, MP, HRW, JM, EB, IR, JH, CG, AS, PBM, MJS, MP

## Data and materials availability

Our Future Health data cannot be shared by the authors but are available to approved researchers via the Our Future Health Trusted Research Environment; access conditions are described at https://research.ourfuturehealth.org.uk/apply-to-access-the-data/. This study was conducted under study ID OFHS240171. All outputs underlying the findings are aggregate summaries available in the main text and supplementary materials. Analysis code is available at https://github.com/ehr-lshtm/ofh-pharmcat-755k.

## References

1. M. Ingelman-Sundberg, C. Rodriguez-Antona, Pharmacogenetics of drug-metabolizing enzymes: implications for a safer and more effective drug therapy. Philos. Trans. R. Soc. Lond. B. Biol. Sci. 360, 1563–1570 (2005).

2. S. Ahmed, Z. Zhou, J. Zhou, S.-Q. Chen, Pharmacogenomics of Drug Metabolizing Enzymes and Transporters: Relevance to Precision Medicine. Genomics Proteomics Bioinformatics 14, 298–313 (2016).

3. R. Osanlou, L. Walker, D. A. Hughes, G. Burnside, M. Pirmohamed, Adverse drug reactions, multimorbidity and polypharmacy: a prospective analysis of 1 month of medical admissions. BMJ Open 12, e055551 (2022).

4. E. F. Magavern, M. Megase, J. Thompson, G. Marengo, J. Jacobsen, D. Smedley, M. J. Caulfield, Pharmacogenetics and adverse drug reports: Insights from a United Kingdom national pharmacovigilance database. PLOS Med. 22, e1004565 (2025).

5. M. V. Relling, T. E. Klein, CPIC: Clinical Pharmacogenetics Implementation Consortium of the Pharmacogenomics Research Network. Clin. Pharmacol. Ther. 89, 464–467 (2011).

6. K. E. Caudle, M. Whirl Carrillo, M. V. Relling, J. M. Hoffman, R. S. Donnelly, C. E. Haidar, M. S. Bourque, S. Frear, L. Gong, K. Sangkuhl, R. Whaley, T. E. Klein, Advancing Clinical Pharmacogenomics Worldwide Through the Clinical Pharmacogenetics Implementation Consortium (CPIC). Clin. Pharmacol. Ther. 118, 1512–1522 (2025).

7. J. J. Swen, I. Wilting, A. L. de Goede, L. Grandia, H. Mulder, D. J. Touw, A. de Boer, J. M. H. Conemans, T. C. G. Egberts, O. H. Klungel, R. Koopmans, J. van der Weide, B. Wilffert, H.-J. Guchelaar, V. H. M. Deneer, Pharmacogenetics: from bench to byte. Clin. Pharmacol. Ther. 83, 781–787 (2008).

8. J. J. Swen, M. Nijenhuis, A. de Boer, L. Grandia, A. H. Maitland-van der Zee, H. Mulder, G. a. P. J. M. Rongen, R. H. N. van Schaik, T. Schalekamp, D. J. Touw, J. van der Weide, B. Wilffert, V. H. M. Deneer, H.-J. Guchelaar, Pharmacogenetics: from bench to byte--an update of guidelines. Clin. Pharmacol. Ther. 89, 662–673 (2011).

9. M. Pirmohamed, C. Dello Russo, UK Centre of Excellence in Regulatory Science and Innovation in Pharmacogenomics. Br. J. Clin. Pharmacol. 92, 327–328 (2026).

10. J. J. Swen, C. H. van der Wouden, L. E. Manson, H. Abdullah-Koolmees, K. Blagec, T. Blagus, S. Böhringer, A. Cambon-Thomsen, E. Cecchin, K.-C. Cheung, V. H. Deneer, M. Dupui, M. Ingelman-Sundberg, S. Jonsson, C. Joefield-Roka, K. S. Just, M. O. Karlsson, L. Konta, R. Koopmann, M. Kriek, T. Lehr, C. Mitropoulou, E. Rial-Sebbag, V. Rollinson, R. Roncato, M. Samwald, E. Schaeffeler, M. Skokou, M. Schwab, D. Steinberger, J. C. Stingl, R. Tremmel, R. M. Turner, M. H. van Rhenen, C. L. Dávila Fajardo, V. Dolžan, G. P. Patrinos, M. Pirmohamed, G. Sunder-Plassmann, G. Toffoli, H.-J. Guchelaar, Ubiquitous Pharmacogenomics Consortium, A 12-gene pharmacogenetic panel to prevent adverse drug reactions: an open-label, multicentre, controlled, cluster-randomised crossover implementation study. Lancet 401, 347–356 (2023).

11. NICE, CYP2C19 genotype testing to guide clopidogrel use after ischaemic stroke or transient ischaemic attack (2024; https://www.nice.org.uk/guidance/htg724).

12. C. Dello Russo, I. Frater, R. Kuruvilla, S. Lip, H. O’Neill, K. Burke, V. Chaplin, A. S. F. Doney, S. Elyas, N. Greaves, S. Harding, D. Hargroves, J. Hayward, D. A. Hughes, T. A. T. Hughes, S. Kondapally, P. Mok, A. Peace, I. Rafi, S. Ray, V. Stinton, L. Venetucci, M. Pirmohamed, CYP2C19 genotype testing for clopidogrel: A guideline developed by the UK Centre of Excellence in Regulatory Science and Innovation in Pharmacogenomics (CERSI-PGx). Br. J. Clin. Pharmacol. 92, 329–347 (2026).

13. J. H. McDermott, M. Tsakiroglou, W. G. Newman, M. Pirmohamed, Pharmacogenomics in the UK National Health Service: Progress towards implementation. Br. J. Clin. Pharmacol. 91, 2241–2250 (2025).

14. G. McInnes, A. Lavertu, K. Sangkuhl, T. E. Klein, M. Whirl Carrillo, R. B. Altman, Pharmacogenetics at Scale: An Analysis of the UK Biobank. Clin. Pharmacol. Ther. 109, 1528–1537 (2021).

15. A. Haddad, A. Radhakrishnan, S. McGee, J. D. Smith, J. H. Karnes, E. Venner, M. M. Wheeler, K. Patterson, K. Walker, D. Kalra, S. E. Kalla, Q. Wang, R. A. Gibbs, G. P. Jarvik, J. Sanchez, A. Musick, A. H. Ramirez, J. C. Denny, P. E. Empey, Frequency of pharmacogenomic variation and medication exposures among All of Us Participants (2024), doi:10.1101/2024.06.12.24304664.

16. S. S. Verma, K. Keat, B. Li, G. Hoffecker, M. Risman, Regeneron Genetics Center, K. Sangkuhl, M. Whirl-Carrillo, S. Dudek, A. Verma, T. E. Klein, M. D. Ritchie, S. Tuteja, Evaluating the frequency and the impact of pharmacogenetic alleles in an ancestrally diverse Biobank population. J. Transl. Med. 20, 550 (2022).

17. Y. Zhou, M. Ingelman-Sundberg, V. M. Lauschke, Worldwide Distribution of Cytochrome P450 Alleles: A Meta-analysis of Population-scale Sequencing Projects. Clin. Pharmacol. Ther. 102, 688–700 (2017).

18. C. Chanfreau-Coffinier, L. E. Hull, J. A. Lynch, S. L. DuVall, S. M. Damrauer, F. E. Cunningham, B. F. Voight, M. E. Matheny, D. W. Oslin, M. S. Icardi, S. Tuteja, Projected Prevalence of Actionable Pharmacogenetic Variants and Level A Drugs Prescribed Among US Veterans Health Administration Pharmacy Users. *JAMA Netw*. Open 2, e195345 (2019).

19. M. B. Cook, N. Adams, A. Adjetey, R. Arathimos, M. Balabanovic, R. Blackwood, A. Booth, B. J. Cairns, A. Connell, S. Ellis, B. Elsworth, K. Evans, A. Forman, E. Gradovich, C. Gretton, F. Grimm, D. J. Hunter, K. Lipinski, J. Lord, J. Luff, F. Maleady-Crowe, R. Moran, S. North, A. Peel, D. Van Der Plaat, K. Purves, F. Reddington, A. Roddam, S. C. Sanderson, T. Sprosen, A. Steventon, I. Turnbull, E. Vestesson, R. Ali, Cohort Profile: Our Future Health. Int. J. Epidemiol. 54, dyaf171 (2025).

20. P. Dixon, W. G. Newman, V. Sharma, J. H. McDermott, C. Wright Drakesmith, Self-Reported Pharmacogenetic Medication Use in the Our Future Health Cohort. Clin. Transl. Sci. 19, e70471 (2026).

21. N. Daneshi, E. Holliday, S. Hancock, J. J. Schneider, R. J. Scott, J. Attia, E. A. Milward, Prevalence of clinically actionable genotypes and medication exposure of older adults in the community. Pharmacogenomics Pers. Med. 10, 17–27 (2017).

22. M. Pirmohamed, Pharmacogenomics: current status and future perspectives. Nat. Rev. Genet. 24, 350–362 (2023).

23. National Academies of Sciences, Engineering, and Medicine; Division of Behavioral and Social Sciences and Education; Health and Medicine Division; Committee on Population; Board on Health Sciences Policy; Committee on the Use of Race, Ethnicity, and Ancestry as Population Descriptors in Genomics Research, Using Population Descriptors in Genetics and Genomics Research: A New Framework for an Evolving Field (National Academies Press (US), Washington (DC), 2023; http://www.ncbi.nlm.nih.gov/books/NBK589855/).

24. J. A. Johnson, K. E. Caudle, L. Gong, M. Whirl-Carrillo, C. M. Stein, S. A. Scott, M. T. Lee, B. F. Gage, S. E. Kimmel, M. A. Perera, J. L. Anderson, M. Pirmohamed, T. E. Klein, N. A. Limdi, L. H. Cavallari, M. Wadelius, Clinical Pharmacogenetics Implementation Consortium (CPIC) Guideline for Pharmacogenetics-Guided Warfarin Dosing: 2017 Update. Clin. Pharmacol. Ther. 102, 397–404 (2017).

25. J. P. Jarvis, A. P. Peter, J. A. Shaman, Consequences of CYP2D6 Copy-Number Variation for Pharmacogenomics in Psychiatry. Front. Psychiatry 10, 432 (2019).

26. PharmCAT, Calling CYP2D6 (available at https://pharmcat.clinpgx.org/using/Calling-CYP2D6/).

27. PharmCAT, Calling HLA-A and B Alleles (available at https://pharmcat.clinpgx.org/using/Calling-HLA/).

28. S. Murtough, O. Stellakis, D. Mills, B. Bjourson, V. Chaplin, D. Chauhan, B. Chipp, M. Cotic, J. de Villiers, O. Dzahini, F. Elmslie, K. Evans, S. Gandhi, D. A. Hughes, H. Jin, D. Panconesi, A. Skowronska, S. M. Sisodiya, E. Silva, V. Stinton, S. Stuart-Smith, S. Tarrant, D. Taylor, L. Varney, J. T. R. Walters, M. Wood, J. Woodley, C. Dello Russo, M. Pirmohamed, E. Bramon, ACKR1/Duffy-null genotype testing for clozapine: A guideline developed by the UK Centre of Excellence in Regulatory Science and Innovation in Pharmacogenomics (CERSI-PGx). Br. J. Clin. Pharmacol. 92, 1977–1990 (2026).

29. Royal College of Physicians and British Pharmacological Society, Personalised prescribing: using pharmacogenomics to improve patient outcomes (London: RCP and BPS, 2022; https://www.bps.ac.uk/fileadmin/uploads/bps/Reports/Personalised-prescribing_main_report_2022.pdf).

30. European Medicines Agency, Multi-stakeholder workshop on pharmacogenomics (EMA: Amsterdam, 2024; https://www.ema.europa.eu/en/documents/report/report-joint-ec-hma-ema-multi-stakeholder-workshop-pharmacogenomics-24-september-2024_en.pdf).

31. Our Future Health, Imputed genotype data (available at https://ourfuturehealth.gitbook.io/our-future-health/data-types/genetic-data/imputed-genotype-data).

32. T. E. Klein, M. D. Ritchie, PharmCAT: A Pharmacogenomics Clinical Annotation Tool. Clin. Pharmacol. Ther. 104, 19–22 (2018).

33. K. Sangkuhl, M. Whirl-Carrillo, R. M. Whaley, M. Woon, A. Lavertu, R. B. Altman, L. Carter, A. Verma, M. D. Ritchie, T. E. Klein, Pharmacogenomics Clinical Annotation Tool (PharmCAT). Clin. Pharmacol. Ther. 107, 203–210 (2020).

34. PharmCAT v3.1.1 (2025) (available at https://github.com/PharmGKB/PharmCAT/releases/tag/v3.1.1).

35. R. M. Cooper-DeHoff, M. Niemi, L. B. Ramsey, J. A. Luzum, E. K. Tarkiainen, R. J. Straka, L. Gong, S. Tuteja, R. A. Wilke, M. Wadelius, E. A. Larson, D. M. Roden, T. E. Klein, S. W. Yee, R. M. Krauss, R. M. Turner, L. Palaniappan, A. Gaedigk, K. M. Giacomini, K. E. Caudle, D. Voora, The Clinical Pharmacogenetics Implementation Consortium Guideline for SLCO1B1, ABCG2, and CYP2C9 genotypes and Statin-Associated Musculoskeletal Symptoms. Clin. Pharmacol. Ther. 111, 1007–1021 (2022).

36. Our Future Health, Genetic ancestry (available at https://ourfuturehealth.gitbook.io/our-future-health/data-types/genetic-data/genetic-ancestry).

37. M. Whirl-Carrillo, R. Huddart, L. Gong, K. Sangkuhl, C. F. Thorn, R. Whaley, T. E. Klein, An Evidence-Based Framework for Evaluating Pharmacogenomics Knowledge for Personalized Medicine. Clin. Pharmacol. Ther. 110, 563–572 (2021).

38. Bennett Institute for Applied Data Science, OpenPrescribing.net (2026) (available at https://openprescribing.net).

