## Supplement for "Genotype-predicted drug response phenotypes and their co-occurrence with dispensed medicines among 738,531 participants in the UK Our Future Health study"

| Table S1. Mapping between genetically predicted drug response phenotypes and actionability | | | |
| --- | --- | --- | --- |
|  | **Phenotype** | **Actionability** | **n (%)** |
| *ABCG2* | Decreased Function | Actionable | 143,966 (19.5) |
|  | Poor Function | Actionable | 9,655 (1.3) |
|  | Normal Function | Not Actionable | 584,910 (79.2) |
| *CACNA1S* | Malignant Hyperthermia Susceptibility | Actionable | redacted |
|  | Uncertain Susceptibility | Not Actionable | redacted |
| *CFTR* | Non-responsive | Actionable | 649,189 (87.9) |
|  | ivacaftor responsive in CF patients | Actionable | 75,593 (10.2) |
|  | Indeterminate/Missing | Indeterminate/Missing | 13,749 (1.9) |
| *CYP2B6* | Intermediate Metabolizer | Actionable | 254,748 (34.5) |
|  | Poor Metabolizer | Actionable | 46,068 (6.2) |
|  | Indeterminate/Missing | Indeterminate/Missing | 58,994 (8.0) |
|  | Normal Metabolizer | Not Actionable | 378,721 (51.3) |
| *CYP2C19* | Intermediate Metabolizer | Actionable | 201,629 (27.3) |
|  | Rapid Metabolizer | Actionable | 191,581 (25.9) |
|  | Ultrarapid Metabolizer | Actionable | 32,983 (4.5) |
|  | Poor Metabolizer | Actionable | 22,092 (3.0) |
|  | Indeterminate/Missing | Indeterminate/Missing | 1,163 (0.2) |
|  | Normal Metabolizer | Not Actionable | 289,083 (39.1) |
| *CYP2C9* | Intermediate Metabolizer | Actionable | 163,255 (22.1) |
|  | Poor Metabolizer | Actionable | 192 (0.0) |
|  | Indeterminate/Missing | Indeterminate/Missing | 128,023 (17.3) |
|  | Normal Metabolizer | Not Actionable | 447,061 (60.5) |
| *CYP3A4* | Intermediate Metabolizer | Actionable | 59,274 (8.0) |
|  | Poor Metabolizer | Actionable | 1,607 (0.2) |
|  | Indeterminate/Missing | Indeterminate/Missing | 18,563 (2.5) |
|  | Normal Metabolizer | Not Actionable | 659,087 (89.2) |
| *CYP3A5* | Poor Metabolizer | Actionable | 616,815 (83.5) |
|  | Intermediate Metabolizer | Actionable | 111,206 (15.1) |
|  | Indeterminate/Missing | Indeterminate/Missing | redacted |
|  | Normal Metabolizer | Not Actionable | redacted |
| *DPYD* | Intermediate Metabolizer | Actionable | 46,456 (6.3) |
|  | Poor Metabolizer | Actionable | 199 (0.0) |
|  | Normal Metabolizer | Not Actionable | 691,876 (93.7) |
| *G6PD* | Variable | Actionable | 3,480 (0.5) |
|  | Deficient | Actionable | 737 (0.1) |
|  | Indeterminate/Missing | Indeterminate/Missing | 5,728 (0.8) |
|  | Normal | Not Actionable | 728,586 (98.7) |
| *NAT2* | Poor Metabolizer | Actionable | 428,749 (58.1) |
|  | Intermediate Metabolizer | Actionable | 8,552 (1.2) |
|  | Rapid Metabolizer | Actionable | 107 (0.0) |
|  | Indeterminate/Missing | Indeterminate/Missing | 301,123 (40.8) |
| *NUDT15* | Intermediate Metabolizer | Actionable | 12,825 (1.7) |
|  | Poor Metabolizer | Actionable | 256 (0.0) |
|  | Indeterminate/Missing | Indeterminate/Missing | 6,650 (0.9) |
|  | Normal Metabolizer | Not Actionable | 718,800 (97.3) |
| *RYR1* | Malignant Hyperthermia Susceptibility | Actionable | 127 (0.0) |
|  | Uncertain Susceptibility | Not Actionable | 738,404 (100.0) |
| *SLCO1B1* | Decreased Function | Actionable | 183,073 (24.8) |
|  | Poor Function | Actionable | 16,612 (2.2) |
|  | Possible Decreased Function | Actionable | 563 (0.1) |
|  | Indeterminate/Missing | Indeterminate/Missing | 8,552 (1.2) |
|  | Normal Function | Not Actionable | 499,203 (67.6) |
|  | Increased Function | Not Actionable | 30,528 (4.1) |
| *TPMT* | Intermediate Metabolizer | Actionable | 66,994 (9.1) |
|  | Poor Metabolizer | Actionable | 1,837 (0.2) |
|  | Indeterminate/Missing | Indeterminate/Missing | 2,365 (0.3) |
|  | Normal Metabolizer | Not Actionable | 667,335 (90.4) |
| *UGT1A1* | Poor Metabolizer | Actionable | 725,038 (98.2) |
|  | Intermediate Metabolizer | Actionable | 3,274 (0.4) |
|  | Indeterminate/Missing | Indeterminate/Missing | 6,833 (0.9) |
|  | Normal Metabolizer | Not Actionable | 3,386 (0.5) |
| *VKORC1* | -1639 AG | Actionable | 334,487 (45.3) |
|  | -1639 AA | Actionable | 111,294 (15.1) |
|  | -1639 GG | Not Actionable | 292,750 (39.6) |
| *Notes:* Genetically predicted drug response phenotypes were assigned by Pharmacogenomics Clinical Annotation Tool (PharmCAT v3.1.1) and classified as actionable when guidelines would advise deviating from standard care. Percentages are within gene. Cells are marked redacted to protect small cell counts; more than one cell may be redacted within a gene for this purpose, so a redacted cell does not always indicate a small cell count. Two genes (*CYP4F2* and *IFNL3*) had no assigned phenotype and were excluded from all analyses. PharmCAT assigns no “Normal Metabolizer” phenotype for *NAT2*; all participants with a determinate call are therefore actionable. | | | |

| Table S2. Prevalence and 95% confidence intervals of actionable pharmacogenomic phenotypes by genetically-inferred ancestry | | | | | | |
| --- | --- | --- | --- | --- | --- | --- |
|  | **EUR (n=669,368)** | **SAS (n=33,935)** | **EAS (n=13,906)** | **AFR (n=11,379)** | **MENA (n=8,341)** | **AMR (n=1,602)** |
| *ABCG2* | 20.7 (20.6 to 20.8) | 18.1 (17.7 to 18.5) | 49.1 (48.3 to 50.0) | 2.8 (2.5 to 3.1) | 11.6 (10.9 to 12.3) | 31.5 (29.3 to 33.8) |
| *CFTR* | 98.0 (98.0 to 98.1) | 99.7 (99.7 to 99.8) | 100.0 (99.9 to 100.0) | 97.3 (97.0 to 97.6) | 98.6 (98.3 to 98.8) | 98.4 (97.6 to 98.9) |
| *CYP2B6* | 39.4 (39.3 to 39.5) | 57.3 (56.7 to 57.8) | 40.0 (39.2 to 40.9) | 62.6 (61.7 to 63.5) | 47.1 (46.0 to 48.1) | 55.2 (52.7 to 57.6) |
| *CYP2C19* | 60.1 (60.0 to 60.2) | 73.7 (73.3 to 74.2) | 61.0 (60.2 to 61.8) | 64.5 (63.7 to 65.4) | 57.1 (56.0 to 58.2) | 37.3 (35.0 to 39.7) |
| *CYP2C9* | 23.1 (23.0 to 23.2) | 11.6 (11.3 to 12.0) | 1.7 (1.5 to 1.9) | 22.5 (21.7 to 23.2) | 20.9 (20.1 to 21.8) | 14.5 (12.9 to 16.4) |
| *CYP3A4* | 8.8 (8.7 to 8.8) | 2.3 (2.2 to 2.5) | 4.5 (4.2 to 4.9) | 1.3 (1.1 to 1.6) | 6.2 (5.7 to 6.7) | 3.7 (2.9 to 4.7) |
| *CYP3A5* | 99.5 (99.5 to 99.6) | 92.5 (92.2 to 92.8) | 92.4 (92.0 to 92.8) | 68.2 (67.4 to 69.1) | 98.1 (97.7 to 98.3) | 97.2 (96.3 to 97.9) |
| *DPYD* | 6.6 (6.5 to 6.6) | 5.3 (5.0 to 5.5) | 0.4 (0.3 to 0.6) | 4.9 (4.5 to 5.3) | 4.0 (3.6 to 4.5) | 2.5 (1.8 to 3.4) |
| *G6PD* | 0.2 (0.2 to 0.2) | 2.7 (2.6 to 2.9) | 2.9 (2.7 to 3.2) | 9.7 (9.2 to 10.3) | 6.0 (5.5 to 6.5) | 1.1 (0.7 to 1.7) |
| *NAT2* | 59.9 (59.8 to 60.0) | 60.8 (60.3 to 61.3) | 24.0 (23.3 to 24.7) | 59.3 (58.4 to 60.2) | 61.3 (60.2 to 62.3) | 43.4 (41.0 to 45.9) |
| *NUDT15* | 0.9 (0.9 to 0.9) | 12.4 (12.1 to 12.8) | 17.3 (16.6 to 17.9) | 0.4 (0.3 to 0.5) | 2.5 (2.2 to 2.9) | 11.2 (9.7 to 12.8) |
| *SLCO1B1* | 28.2 (28.1 to 28.3) | 10.4 (10.1 to 10.7) | 23.6 (22.9 to 24.3) | 17.1 (16.4 to 17.8) | 28.4 (27.5 to 29.4) | 27.7 (25.6 to 30.0) |
| *TPMT* | 9.8 (9.7 to 9.8) | 3.5 (3.4 to 3.7) | 3.5 (3.2 to 3.9) | 10.1 (9.6 to 10.7) | 4.5 (4.1 to 5.0) | 10.0 (8.6 to 11.6) |
| *UGT1A1* | 99.4 (99.4 to 99.4) | 94.9 (94.6 to 95.1) | 70.2 (69.4 to 71.0) | 99.7 (99.6 to 99.8) | 98.4 (98.1 to 98.7) | 96.1 (95.1 to 97.0) |
| *VKORC1* | 61.5 (61.4 to 61.7) | 35.9 (35.4 to 36.5) | 97.8 (97.5 to 98.0) | 15.1 (14.4 to 15.7) | 62.2 (61.2 to 63.3) | 72.3 (70.0 to 74.4) |
| *Abbreviations:* EUR, European ancestry; SAS, South Asian ancestry; EAS, East Asian ancestry; AFR, African ancestry; MENA, Middle Eastern or North African ancestry; AMR, Admixed American ancestry | | | | | | |
| *Notes:* Genetically predicated drug response phenotypes were assigned by Pharmacogenomics Clinical Annotation Tool (PharmCAT v3.1.1) and classified as actionable when guidelines would advise deviating from standard care. Percentages are within ancestry group. Two genes (*CYP4F2* and *IFNL3*) had no assigned phenotype and were excluded from all analyses. Two additional genes (*CACNA1S* and *RYR1*) were excluded due to small cell sizes in all ancestry groups. 95% confidence intervals were calculated using the Wilson score method without continuity correction. | | | | | | |

| Table S3. Distribution of genetically predicted drug response phenotypes by genetically-inferred ancestry | | | | | | | |
| --- | --- | --- | --- | --- | --- | --- | --- |
|  | **Phenotype** | **EUR (n=669,368)** | **SAS (n=33,935)** | **EAS (n=13,906)** | **AFR (n=11,379)** | **MENA (n=8,341)** | **AMR (n=1,602)** |
| *ABCG2* | Decreased Function | 130,788 (19.5) | 5,788 (17.1) | 5,677 (40.8) | redacted | 935 (11.2) | 463 (28.9) |
|  | Normal Function | 530,503 (79.3) | 27,798 (81.9) | 7,074 (50.9) | 11,062 (97.2) | 7,376 (88.4) | 1,097 (68.5) |
|  | Poor Function | 8,077 (1.2) | 349 (1.0) | 1,155 (8.3) | redacted | redacted | 42 (2.6) |
| *CFTR* | Indeterminate/Missing | 13,198 (2.0) | 94 (0.3) | redacted | 306 (2.7) | 120 (1.4) | redacted |
|  | Non-responsive | 583,131 (87.1) | 32,318 (95.2) | 13,711 (98.6) | 10,904 (95.8) | 7,625 (91.4) | 1,500 (93.6) |
|  | ivacaftor responsive in CF patients | 73,039 (10.9) | 1,523 (4.5) | redacted | 169 (1.5) | 596 (7.1) | 76 (4.7) |
| *CYP2B6* | Indeterminate/Missing | 52,736 (7.9) | 2,426 (7.1) | 672 (4.8) | 2,018 (17.7) | 1,008 (12.1) | 134 (8.4) |
|  | Intermediate Metabolizer | 226,890 (33.9) | 14,600 (43.0) | 4,794 (34.5) | 4,608 (40.5) | 3,180 (38.1) | 676 (42.2) |
|  | Normal Metabolizer | 352,754 (52.7) | 12,075 (35.6) | 7,666 (55.1) | 2,235 (19.6) | 3,407 (40.8) | 584 (36.5) |
|  | Poor Metabolizer | 36,988 (5.5) | 4,834 (14.2) | 774 (5.6) | 2,518 (22.1) | 746 (8.9) | 208 (13.0) |
| *CYP2C19* | Indeterminate/Missing | 768 (0.1) | redacted | redacted | 316 (2.8) | 50 (0.6) | redacted |
|  | Intermediate Metabolizer | 174,683 (26.1) | 14,692 (43.3) | 6,475 (46.6) | 3,525 (31.0) | 1,953 (23.4) | 301 (18.8) |
|  | Normal Metabolizer | 266,527 (39.8) | 8,893 (26.2) | 5,418 (39.0) | 3,719 (32.7) | 3,529 (42.3) | redacted |
|  | Poor Metabolizer | 15,811 (2.4) | 3,753 (11.1) | 1,817 (13.1) | 509 (4.5) | 177 (2.1) | 25 (1.6) |
|  | Rapid Metabolizer | 180,620 (27.0) | 5,517 (16.3) | redacted | 2,781 (24.4) | 2,210 (26.5) | 258 (16.1) |
|  | Ultrarapid Metabolizer | 30,959 (4.6) | 1,058 (3.1) | redacted | 529 (4.6) | 422 (5.1) | redacted |
| *CYP2C9* | Indeterminate/Missing | 117,619 (17.6) | 7,230 (21.3) | 991 (7.1) | 560 (4.9) | redacted | 137 (8.6) |
|  | Intermediate Metabolizer | 154,593 (23.1) | redacted | 230 (1.7) | 2,511 (22.1) | redacted | 233 (14.5) |
|  | Normal Metabolizer | 397,021 (59.3) | 22,752 (67.0) | 12,685 (91.2) | 8,262 (72.6) | 5,109 (61.3) | 1,232 (76.9) |
|  | Poor Metabolizer | 135 (0.0) | redacted | redacted | 46 (0.4) | redacted | redacted |
| *CYP3A4* | Indeterminate/Missing | 17,176 (2.6) | redacted | redacted | 799 (7.0) | redacted | 22 (1.4) |
|  | Intermediate Metabolizer | 57,152 (8.5) | redacted | redacted | 153 (1.3) | redacted | 59 (3.7) |
|  | Normal Metabolizer | 593,454 (88.7) | 32,888 (96.9) | 13,145 (94.5) | 10,427 (91.6) | 7,652 (91.7) | 1,521 (94.9) |
|  | Poor Metabolizer | 1,586 (0.2) | redacted | redacted | redacted | redacted | redacted |
| *CYP3A5* | Indeterminate/Missing | redacted | redacted | redacted | redacted | redacted | redacted |
|  | Intermediate Metabolizer | 84,830 (12.7) | 12,737 (37.5) | 5,651 (40.6) | 5,452 (47.9) | 2,056 (24.6) | 480 (30.0) |
|  | Normal Metabolizer | 3,092 (0.5) | 2,534 (7.5) | 1,054 (7.6) | 3,614 (31.8) | 162 (1.9) | 45 (2.8) |
|  | Poor Metabolizer | 581,440 (86.9) | 18,662 (55.0) | 7,201 (51.8) | 2,312 (20.3) | 6,123 (73.4) | 1,077 (67.2) |
| *DPYD* | Intermediate Metabolizer | 43,686 (6.5) | 1,780 (5.2) | 61 (0.4) | 555 (4.9) | redacted | 40 (2.5) |
|  | Normal Metabolizer | 625,496 (93.4) | 32,144 (94.7) | 13,845 (99.6) | 10,824 (95.1) | 8,005 (96.0) | 1,562 (97.5) |
|  | Poor Metabolizer | 186 (0.0) | redacted | redacted | redacted | redacted | redacted |
| *G6PD* | Deficient | 258 (0.0) | 271 (0.8) | redacted | redacted | 141 (1.7) | redacted |
|  | Indeterminate/Missing | 737 (0.1) | 340 (1.0) | 20 (0.1) | 4,135 (36.3) | 459 (5.5) | 37 (2.3) |
|  | Normal | 667,374 (99.7) | 32,666 (96.3) | 13,477 (96.9) | 6,138 (53.9) | 7,383 (88.5) | 1,548 (96.6) |
|  | Variable | 999 (0.1) | 658 (1.9) | 347 (2.5) | redacted | 358 (4.3) | 17 (1.1) |
| *NAT2* | Indeterminate/Missing | 268,498 (40.1) | 13,293 (39.2) | 10,568 (76.0) | 4,628 (40.7) | 3,230 (38.7) | 906 (56.6) |
|  | Intermediate Metabolizer | 6,302 (0.9) | redacted | redacted | 1,254 (11.0) | redacted | 33 (2.1) |
|  | Poor Metabolizer | 394,553 (58.9) | 19,941 (58.8) | 3,277 (23.6) | 5,419 (47.6) | 4,896 (58.7) | 663 (41.4) |
|  | Rapid Metabolizer | 15 (0.0) | redacted | redacted | 78 (0.7) | redacted | redacted |
| *NUDT15* | Indeterminate/Missing | 5,739 (0.9) | 187 (0.6) | 578 (4.2) | 72 (0.6) | redacted | redacted |
|  | Intermediate Metabolizer | 6,017 (0.9) | 4,095 (12.1) | 2,288 (16.5) | 43 (0.4) | redacted | redacted |
|  | Normal Metabolizer | 657,596 (98.2) | 29,530 (87.0) | 10,928 (78.6) | 11,264 (99.0) | 8,083 (96.9) | 1,399 (87.3) |
|  | Poor Metabolizer | redacted | 123 (0.4) | 112 (0.8) | redacted | redacted | redacted |
| *SLCO1B1* | Decreased Function | 172,467 (25.8) | 3,397 (10.0) | 3,046 (21.9) | 1,644 (14.4) | 2,119 (25.4) | 400 (25.0) |
|  | Increased Function | 30,051 (4.5) | redacted | redacted | 95 (0.8) | 233 (2.8) | redacted |
|  | Indeterminate/Missing | 3,404 (0.5) | 2,401 (7.1) | 198 (1.4) | 2,180 (19.2) | 347 (4.2) | 22 (1.4) |
|  | Normal Function | 447,238 (66.8) | 27,870 (82.1) | 10,432 (75.0) | 7,157 (62.9) | 5,390 (64.6) | 1,116 (69.7) |
|  | Poor Function | 15,900 (2.4) | redacted | 200 (1.4) | 117 (1.0) | 220 (2.6) | redacted |
|  | Possible Decreased Function | 308 (0.0) | redacted | redacted | 186 (1.6) | 32 (0.4) | redacted |
| *TPMT* | Indeterminate/Missing | 1,137 (0.2) | 41 (0.1) | redacted | 976 (8.6) | redacted | redacted |
|  | Intermediate Metabolizer | 63,679 (9.5) | 1,182 (3.5) | redacted | 1,118 (9.8) | redacted | redacted |
|  | Normal Metabolizer | 602,785 (90.1) | 32,692 (96.3) | 13,354 (96.0) | 9,249 (81.3) | 7,822 (93.8) | redacted |
|  | Poor Metabolizer | 1,767 (0.3) | 20 (0.1) | redacted | 36 (0.3) | redacted | redacted |
| *UGT1A1* | Indeterminate/Missing | 962 (0.1) | 1,619 (4.8) | 4,073 (29.3) | redacted | 116 (1.4) | redacted |
|  | Intermediate Metabolizer | 2,995 (0.4) | 144 (0.4) | 44 (0.3) | 43 (0.4) | 38 (0.5) | redacted |
|  | Normal Metabolizer | 3,149 (0.5) | 122 (0.4) | 69 (0.5) | redacted | 17 (0.2) | redacted |
|  | Poor Metabolizer | 662,262 (98.9) | 32,050 (94.4) | 9,720 (69.9) | 11,306 (99.4) | 8,170 (97.9) | 1,530 (95.5) |
| *VKORC1* | -1639 AA | 97,137 (14.5) | 1,796 (5.3) | 10,505 (75.5) | 167 (1.5) | 1,320 (15.8) | 369 (23.0) |
|  | -1639 AG | 314,789 (47.0) | 10,401 (30.6) | 3,091 (22.2) | 1,547 (13.6) | 3,870 (46.4) | 789 (49.3) |
|  | -1639 GG | 257,442 (38.5) | 21,738 (64.1) | 310 (2.2) | 9,665 (84.9) | 3,151 (37.8) | 444 (27.7) |
| *Abbreviations:* EUR, European ancestry; SAS, South Asian ancestry; EAS, East Asian ancestry; AFR, African ancestry; MENA, Middle Eastern or North African ancestry; AMR, Admixed American ancestry | | | | | | | |
| *Notes:* Genetically predicated drug response phenotypes were assigned by Pharmacogenomics Clinical Annotation Tool (PharmCAT v3.1.1) and classified as actionable when guidelines would advise deviating from standard care. Percentages are within gene and ancestry group. Cells are marked redacted to protect small cell counts; more than one cell may be redacted within a gene for this purpose, so a redacted cell does not always indicate a small cell count. Two genes (*CYP4F2* and *IFNL3*) had no assigned phenotype and were excluded from all analyses. Two additional genes (*CACNA1S* and *RYR1*) were excluded due to small cell sizes in all ancestry groups. PharmCAT assigns no “Normal Metabolizer” phenotype for *NAT2*; all participants with a determinate call are therefore actionable. | | | | | | | |

| Table S4. Percentage and 95% confidence intervals of participants dispensed at least one medicine with a guideline-backed recommendation at the paired gene, irrespective of their drug response phenotype | | | | | | |
| --- | --- | --- | --- | --- | --- | --- |
|  | **EUR (n=669,368)** | **SAS (n=33,935)** | **EAS (n=13,906)** | **AFR (n=11,379)** | **MENA (n=8,341)** | **AMR (n=1,602)** |
| *ABCG2* | 5.1 (5.1 to 5.2) | 4.2 (4.0 to 4.4) | 3.3 (3.0 to 3.6) | 3.0 (2.7 to 3.3) | 3.7 (3.3 to 4.1) | 1.4 (0.9 to 2.1) |
| *CFTR* | n/a | n/a | n/a | n/a | n/a | n/a |
| *CYP2B6* | 10.5 (10.5 to 10.6) | 6.5 (6.3 to 6.8) | 3.8 (3.5 to 4.1) | 6.3 (5.9 to 6.8) | 12.3 (11.6 to 13.0) | 8.6 (7.3 to 10.1) |
| *CYP2C19* | 50.6 (50.5 to 50.7) | 38.9 (38.3 to 39.4) | 25.1 (24.4 to 25.9) | 41.2 (40.3 to 42.1) | 47.5 (46.4 to 48.5) | 39.6 (37.2 to 42.0) |
| *CYP2C9* | 3.1 (3.1 to 3.2) | 2.4 (2.2 to 2.5) | 1.1 (0.9 to 1.3) | 2.7 (2.4 to 3.0) | 3.1 (2.8 to 3.5) | 1.5 (1.0 to 2.2) |
| *CYP3A4* | 0.4 (0.4 to 0.5) | 0.3 (0.2 to 0.3) | 0.2 (0.1 to 0.3) | 0.4 (0.3 to 0.5) | 0.7 (0.6 to 1.0) | redacted |
| *CYP3A5* | 0.0 (0.0 to 0.0) | redacted | redacted | redacted | redacted | 0.0 (0.0 to 0.2) |
| *DPYD* | n/a | n/a | n/a | n/a | n/a | n/a |
| *G6PD* | 19.4 (19.3 to 19.5) | 13.9 (13.5 to 14.2) | 9.4 (9.0 to 9.9) | 13.2 (12.6 to 13.8) | 16.4 (15.7 to 17.3) | 19.1 (17.3 to 21.1) |
| *NAT2* | 0.0 (0.0 to 0.0) | 0.0 (0.0 to 0.1) | redacted | 0.1 (0.0 to 0.2) | redacted | 0.0 (0.0 to 0.2) |
| *NUDT15* | 0.3 (0.3 to 0.3) | 0.2 (0.2 to 0.3) | redacted | 0.2 (0.1 to 0.3) | 0.3 (0.2 to 0.4) | redacted |
| *SLCO1B1* | 27.3 (27.1 to 27.4) | 20.4 (20.0 to 20.9) | 11.9 (11.4 to 12.5) | 16.6 (16.0 to 17.3) | 17.4 (16.6 to 18.2) | 9.8 (8.4 to 11.4) |
| *TPMT* | 0.3 (0.3 to 0.3) | 0.2 (0.2 to 0.3) | redacted | 0.2 (0.1 to 0.3) | 0.3 (0.2 to 0.4) | redacted |
| *UGT1A1* | n/a | n/a | n/a | n/a | n/a | n/a |
| *VKORC1* | 0.6 (0.5 to 0.6) | 0.2 (0.1 to 0.2) | 0.1 (0.1 to 0.2) | 0.2 (0.2 to 0.3) | 0.3 (0.2 to 0.5) | redacted |
| *Abbreviations:* EUR, European ancestry; SAS, South Asian ancestry; EAS, East Asian ancestry; AFR, African ancestry; MENA, Middle Eastern or North African ancestry; AMR, Admixed American ancestry; n/a, not applicable due to the gene having no paired medicine prescribed in primary care | | | | | | |
| *Notes:* Dispensing is drawn from linked NHS Business Services Authority primary care records covering April 2018 to June 2025. A participant dispensed more than one medicine paired with the same gene is counted once. A participant can be counted across multiple genes. The set of medicines include 39 gene-drug pairs comprising 33 distinct medicines dispensed in primary care. Two genes (*CYP4F2* and *IFNL3*) had no assigned phenotype and were excluded from all analyses. Two additional genes (*CACNA1S* and *RYR1*) were excluded due to small cell sizes in all ancestry groups. Three genes (*CFTR*, *DPYD*, and *UGT1A1*) have no paired medicine dispensed in primary care. Cells are marked redacted to protect small cell counts; more than one cell may be redacted within a gene for this purpose, so a redacted cell does not always indicate a small cell count. 95% confidence intervals were calculated using the Wilson score method without continuity correction. | | | | | | |

| Table S5. Mapping between sub-continental ancestry regions to genetically-inferred ancestry (GIA) groups | | |
| --- | --- | --- |
| **GIA group** | **Label** | **Region** |
| European (EUR) | *C_S_UK* | Central and southern UK |
|  | *NI_N_SCOT* | Northern Ireland and northern Scotland |
|  | *NW_WALES* | North-west Wales |
|  | *SW_WALES* | South-west Wales |
|  | *N_ENG_S_SCOT* | Northern England and southern Scotland |
|  | *IRELAND* | Ireland |
|  | *FINLAND* | Finland |
|  | *N_EUROPE* | Northern Europe |
|  | *E_EUROPE* | Eastern Europe |
|  | *CW_EUROPE* | Central and western Europe |
|  | *SE_EUROPE* | South-eastern Europe |
|  | *PORTUGAL_SPAIN* | Portugal and Spain |
| South Asian (SAS) | *INDIA_PAKISTAN* | India and Pakistan |
|  | *BANGLADESH* | Bangladesh |
|  | *SRI_LANKA* | Sri Lanka |
|  | *C_ASIA* | Central Asia |
| East Asian (EAS) | *CE_ASIA* | Central-east Asia |
|  | *JAPAN_KOREA* | Japan and Korea |
|  | *SE_ASIA* | South-east Asia |
| African (AFR) | *W_AFRICA* | West Africa |
|  | *E_AFRICA* | East Africa |
|  | *C_S_AFRICA* | Central and southern Africa |
| Middle Eastern and North African (MENA) | *M_EAST_W_ASIA* | Middle East and western Asia |
|  | *N_AFRICA* | North Africa |
| Admixed American (AMR) | *N_C_S_AMERICA* | North, Central and South America |
| *Notes:* Sub-continental ancestry regions computed centrally by Our Future Health and provided to researchers. Further details provided in the Our Future Health documentation at <https://ourfuturehealth.gitbook.io/our-future-health/data-types/genetic-data/genetic-ancestry> | | |

| Table S6. List of all 80 gene-drug pairs associated with 17 of the 19 pharmacogenes covered in PharmCAT v3.1.1 with actionable prescribing guidelines | | | | | | | |
| --- | --- | --- | --- | --- | --- | --- | --- |
| **Gene** | **Drug** | **Recommending body** | **Captured in primary care** | **BNF Chapter** | **BNF Section** | **BNF Paragraph** | **BNF Chemical Code(s)** |
| *ABCG2* | allopurinol | DPWG | yes | 10 Musculoskeletal and Joint Diseases | 10.1 Drugs used in rheumatic diseases and gout | 10.1.4 Gout and cytotoxic induced hyperuricaemia | 1001040C0 |
| *ABCG2* | rosuvastatin | CPIC | yes | 2 Cardiovascular System | 2.12 Lipid-regulating drugs | n/a | 0212000AA |
| *CACNA1S* | desflurane | CPIC | no |  |  |  |  |
| *CACNA1S* | enflurane | CPIC | no |  |  |  |  |
| *CACNA1S* | halothane | CPIC | no |  |  |  |  |
| *CACNA1S* | isoflurane | CPIC | no |  |  |  |  |
| *CACNA1S* | methoxyflurane | CPIC | rare*** |  |  |  |  |
| *CACNA1S* | sevoflurane | CPIC | no |  |  |  |  |
| *CACNA1S* | succinylcholine | CPIC | no |  |  |  |  |
| *CFTR* | ivacaftor | CPIC | rare** |  |  |  |  |
| *CYP2B6* | efavirenz | CPIC & DPWG | rare** |  |  |  |  |
| *CYP2B6* | sertraline | CPIC | yes | 4 Central Nervous System | 4.3 Antidepressant drugs | 4.3.3 Selective serotonin re-uptake inhibitors | 0403030Q0 |
| *CYP2C19* | amitriptyline | CPIC | yes | 4 Central Nervous System | 4.3 Antidepressant drugs | 4.3.1 Tricyclic and related antidepressant drugs | 0403010B0 |
| *CYP2C19* | citalopram | CPIC & DPWG | yes | 4 Central Nervous System | 4.3 Antidepressant drugs | 4.3.3 Selective serotonin re-uptake inhibitors | 0403030D0 / 0403030Z0 |
| *CYP2C19* | clomipramine | CPIC & DPWG | yes | 4 Central Nervous System | 4.3 Antidepressant drugs | 4.3.1 Tricyclic and related antidepressant drugs | 0403010F0 |
| *CYP2C19* | clopidogrel | CPIC & DPWG | yes | 2 Cardiovascular System | 2.9 Antiplatelet drugs | n/a | 0209000C0 |
| *CYP2C19* | dexlansoprazole | CPIC | no |  |  |  |  |
| *CYP2C19* | doxepin | CPIC | yes | 4 Central Nervous System | 4.3 Antidepressant drugs | 4.3.1 Tricyclic and related antidepressant drugs | 0403010L0 |
| *CYP2C19* | escitalopram | CPIC & DPWG | yes | 4 Central Nervous System | 4.3 Antidepressant drugs | 4.3.3 Selective serotonin re-uptake inhibitors | 0403030X0 |
| *CYP2C19* | imipramine | CPIC & DPWG | yes | 4 Central Nervous System | 4.3 Antidepressant drugs | 4.3.1 Tricyclic and related antidepressant drugs | 0403010N0 |
| *CYP2C19* | lansoprazole | CPIC & DPWG | yes | 1 Gastro-Intestinal System | 1.3 Antisecretory drugs and mucosal protectants | 1.3.5 Proton pump inhibitors | 0103050L0 |
| *CYP2C19* | mavacamten | DPWG | no |  |  |  |  |
| *CYP2C19* | omeprazole | CPIC & DPWG | yes | 1 Gastro-Intestinal System | 1.3 Antisecretory drugs and mucosal protectants | 1.3.5 Proton pump inhibitors | 0103050P0 |
| *CYP2C19* | pantoprazole | CPIC & DPWG | yes | 1 Gastro-Intestinal System | 1.3 Antisecretory drugs and mucosal protectants | 1.3.5 Proton pump inhibitors | 0103050R0 |
| *CYP2C19* | sertraline | CPIC & DPWG | yes | 4 Central Nervous System | 4.3 Antidepressant drugs | 4.3.3 Selective serotonin re-uptake inhibitors | 0403030Q0 |
| *CYP2C19* | trimipramine | CPIC | yes | 4 Central Nervous System | 4.3 Antidepressant drugs | 4.3.1 Tricyclic and related antidepressant drugs | 0403010Y0 |
| *CYP2C19* | voriconazole | CPIC & DPWG | rare* |  |  |  |  |
| *CYP2C9* | celecoxib | CPIC | yes | 10 Musculoskeletal and Joint Diseases | 10.1 Drugs used in rheumatic diseases and gout | 10.1.1 Non-steroidal anti-inflammatory drugs | 1001010AH |
| *CYP2C9* | flurbiprofen | CPIC | yes | 10 Musculoskeletal and Joint Diseases | 10.1 Drugs used in rheumatic diseases and gout | 10.1.1 Non-steroidal anti-inflammatory drugs | 1001010I0 |
| *CYP2C9* | fluvastatin | CPIC | yes | 2 Cardiovascular System | 2.12 Lipid-regulating drugs | n/a | 0212000M0 |
| *CYP2C9* | fosphenytoin | CPIC | rare*** |  |  |  |  |
| *CYP2C9* | ibuprofen | CPIC | yes | 10 Musculoskeletal and Joint Diseases | 10.1 Drugs used in rheumatic diseases and gout | 10.1.1 Non-steroidal anti-inflammatory drugs | 1001010J0 / 1001010AD / 1001010AP / 0407010AD |
| *CYP2C9* | lornoxicam | CPIC | no |  |  |  |  |
| *CYP2C9* | meloxicam | CPIC | yes | 10 Musculoskeletal and Joint Diseases | 10.1 Drugs used in rheumatic diseases and gout | 10.1.1 Non-steroidal anti-inflammatory drugs | 1001010AA |
| *CYP2C9* | phenytoin | CPIC & DPWG | yes | 4 Central Nervous System | 4.8 Antiepileptic drugs | 4.8.1 Control of epilepsy | 0408010Q0 / 0408010Z0 / 0408020T0 |
| *CYP2C9* | piroxicam | CPIC | yes | 10 Musculoskeletal and Joint Diseases | 10.1 Drugs used in rheumatic diseases and gout | 10.1.1 Non-steroidal anti-inflammatory drugs | 1001010R0 |
| *CYP2C9* | siponimod | DPWG | rare*** |  |  |  |  |
| *CYP2C9* | tenoxicam | CPIC | rare* |  |  |  |  |
| *CYP2C9* | warfarin | CPIC & DPWG | yes | 2 Cardiovascular System | 2.8 Anticoagulants and protamine | 2.8.2 Oral anticoagulants | 0208020V0 |
| *CYP3A4* | quetiapine | DPWG | yes | 4 Central Nervous System | 4.2 Drugs used in psychoses and related disorders | 4.2.1 Antipsychotic drugs | 0402010AB |
| *CYP3A5* | tacrolimus | CPIC & DPWG | yes | 8 Malignant Disease and Immunosuppression | 8.2 Drugs affecting the immune response | 8.2.2 Corticosteroids and other immunosuppressants | 0802020T0 |
| *DPYD* | capecitabine | CPIC & DPWG | rare** |  |  |  |  |
| *DPYD* | flucytosine | DPWG | no |  |  |  |  |
| *DPYD* | fluorouracil | CPIC & DPWG | no |  |  |  |  |
| *DPYD* | tegafur | DPWG | no |  |  |  |  |
| *G6PD* | dapsone | CPIC | yes | 5 Infections | 5.1 Antibacterial drugs | 5.1.10 Antileprotic drugs | 0501100H0 |
| *G6PD* | methylene blue | CPIC | no |  |  |  |  |
| *G6PD* | nitrofurantoin | CPIC | yes | 5 Infections | 5.1 Antibacterial drugs | 5.1.13 Urinary-tract infections | 0501130R0 |
| *G6PD* | pegloticase | CPIC | no |  |  |  |  |
| *G6PD* | primaquine | CPIC | rare*** |  |  |  |  |
| *G6PD* | rasburicase | CPIC | no |  |  |  |  |
| *G6PD* | tafenoquine | CPIC | no |  |  |  |  |
| *G6PD* | toluidine blue | CPIC | no |  |  |  |  |
| *NAT2* | hydralazine | CPIC | yes | 2 Cardiovascular System | 2.5 Hypertension and heart failure | 2.5.1 Vasodilator antihypertensive drugs | 0205010J0 |
| *NUDT15* | azathioprine | CPIC & DPWG | yes | 8 Malignant Disease and Immunosuppression | 8.2 Drugs affecting the immune response | 8.2.1 Antiproliferative immunosuppressants | 0802010G0 |
| *NUDT15* | mercaptopurine | CPIC & DPWG | yes | 8 Malignant Disease and Immunosuppression | 8.1 Cytotoxic drugs | 8.1.3 Antimetabolites | 0801030L0 |
| *NUDT15* | thioguanine | CPIC & DPWG | rare** |  |  |  |  |
| *RYR1* | desflurane | CPIC | no |  |  |  |  |
| *RYR1* | enflurane | CPIC | no |  |  |  |  |
| *RYR1* | halothane | CPIC | no |  |  |  |  |
| *RYR1* | isoflurane | CPIC | no |  |  |  |  |
| *RYR1* | methoxyflurane | CPIC | rare*** |  |  |  |  |
| *RYR1* | sevoflurane | CPIC | no |  |  |  |  |
| *RYR1* | succinylcholine | CPIC | no |  |  |  |  |
| *SLCO1B1* | atorvastatin | CPIC & DPWG | yes | 2 Cardiovascular System | 2.12 Lipid-regulating drugs | n/a | 0212000B0 |
| *SLCO1B1* | fluvastatin | CPIC | yes | 2 Cardiovascular System | 2.12 Lipid-regulating drugs | n/a | 0212000M0 |
| *SLCO1B1* | lovastatin | CPIC | no |  |  |  |  |
| *SLCO1B1* | pitavastatin | CPIC | no |  |  |  |  |
| *SLCO1B1* | pravastatin | CPIC | yes | 2 Cardiovascular System | 2.12 Lipid-regulating drugs | n/a | 0212000X0 |
| *SLCO1B1* | rosuvastatin | CPIC & DPWG | yes | 2 Cardiovascular System | 2.12 Lipid-regulating drugs | n/a | 0212000AA |
| *SLCO1B1* | simvastatin | CPIC & DPWG | yes | 2 Cardiovascular System | 2.12 Lipid-regulating drugs | n/a | 0212000Y0 / 0212000AC / 0212000AJ |
| *TPMT* | azathioprine | CPIC & DPWG | yes | 8 Malignant Disease and Immunosuppression | 8.2 Drugs affecting the immune response | 8.2.1 Antiproliferative immunosuppressants | 0802010G0 |
| *TPMT* | mercaptopurine | CPIC & DPWG | yes | 8 Malignant Disease and Immunosuppression | 8.1 Cytotoxic drugs | 8.1.3 Antimetabolites | 0801030L0 |
| *TPMT* | thioguanine | CPIC & DPWG | rare** |  |  |  |  |
| *UGT1A1* | atazanavir | CPIC & DPWG | rare*** |  |  |  |  |
| *UGT1A1* | irinotecan | DPWG | no |  |  |  |  |
| *UGT1A1* | sacituzumab govitecan | DPWG | no |  |  |  |  |
| *VKORC1* | acenocoumarol | DPWG | yes | 2 Cardiovascular System | 2.8 Anticoagulants and protamine | 2.8.2 Oral anticoagulants | 0208020H0 |
| *VKORC1* | phenprocoumon | DPWG | rare*** |  |  |  |  |
| *VKORC1* | warfarin | CPIC & DPWG | yes | 2 Cardiovascular System | 2.8 Anticoagulants and protamine | 2.8.2 Oral anticoagulants | 0208020V0 |
| *Abbreviations:* PharmCAT, Pharmacogenomics Clinical Annotation Tool; CPIC, Clinical Pharmacogenetics Implementation Consortium; DPWG, Dutch Pharmacogenetics Working Group; BNF, British National Formulary | | | | | | | |
| *Notes:* "Rare" refers to the number of prescriptions across all primary care practices in England between June 2021 and May 2026: *<1,000 prescriptions; **<100 prescriptions; ***<10 prescriptions. Two of the 19 pharmacogenes covered by PharmCAT v3.1.1 are not represented. *IFNL3* has no CPIC or DPWG prescribing guideline. *CYP4F2* has one guideline drug, warfarin, but PharmCAT v3.1.1 assigned no phenotype for this gene, so actionability was not determined; warfarin remains represented through its pairings with *VKORC1* and *CYP2C9*. | | | | | | | |

STROBE Statement—Checklist of items that should be included in reports of ***cross-sectional studies***

|  | Item No | Recommendation | Page No |
| --- | --- | --- | --- |
| **Title and abstract** | 1 | (*a*) Indicate the study’s design with a commonly used term in the title or the abstract | 2 |
|  |  | (*b*) Provide in the abstract an informative and balanced summary of what was done and what was found | 2 |
| Introduction | | | |
| Background/rationale | 2 | Explain the scientific background and rationale for the investigation being reported | 3-5 |
| Objectives | 3 | State specific objectives, including any prespecified hypotheses | 5 |
| Methods | | | |
| Study design | 4 | Present key elements of study design early in the paper | 15-16 |
| Setting | 5 | Describe the setting, locations, and relevant dates, including periods of recruitment, exposure, follow-up, and data collection | 16-20 |
| Participants | 6 | (*a*) Give the eligibility criteria, and the sources and methods of selection of participants | 16-20 |
| Variables | 7 | Clearly define all outcomes, exposures, predictors, potential confounders, and effect modifiers. Give diagnostic criteria, if applicable | 16-20 |
| Data sources/ measurement | 8* | For each variable of interest, give sources of data and details of methods of assessment (measurement). Describe comparability of assessment methods if there is more than one group | 16-20 |
| Bias | 9 | Describe any efforts to address potential sources of bias | 16-20 |
| Study size | 10 | Explain how the study size was arrived at | 16, 19-20 |
| Quantitative variables | 11 | Explain how quantitative variables were handled in the analyses. If applicable, describe which groupings were chosen and why | 17-18, 20-21 |
| Statistical methods | 12 | (*a*) Describe all statistical methods, including those used to control for confounding | 20-22 |
|  |  | (*b*) Describe any methods used to examine subgroups and interactions | 20-21 |
|  |  | (*c*) Explain how missing data were addressed | 16-18 |
|  |  | (*d*) If applicable, describe analytical methods taking account of sampling strategy | n/a |
|  |  | (*e*) Describe any sensitivity analyses | 21 |
| Results | | | |
| Participants | 13* | (a) Report numbers of individuals at each stage of study—eg numbers potentially eligible, examined for eligibility, confirmed eligible, included in the study, completing follow-up, and analysed | 6 |
|  |  | (b) Give reasons for non-participation at each stage | 6 |
|  |  | (c) Consider use of a flow diagram | n/a |
| Descriptive data | 14* | (a) Give characteristics of study participants (eg demographic, clinical, social) and information on exposures and potential confounders | 6 |
|  |  | (b) Indicate number of participants with missing data for each variable of interest | 6 |
| Outcome data | 15* | Report numbers of outcome events or summary measures | 6-9 |
| Main results | 16 | (*a*) Give unadjusted estimates and, if applicable, confounder-adjusted estimates and their precision (eg, 95% confidence interval). Make clear which confounders were adjusted for and why they were included | 6-9 |
|  |  | (*b*) Report category boundaries when continuous variables were categorized | 8-9 |
|  |  | (*c*) If relevant, consider translating estimates of relative risk into absolute risk for a meaningful time period | 6-9 |
| Other analyses | 17 | Report other analyses done—eg analyses of subgroups and interactions, and sensitivity analyses | 6, 9 |
| Discussion | | | |
| Key results | 18 | Summarise key results with reference to study objectives | 10 |
| Limitations | 19 | Discuss limitations of the study, taking into account sources of potential bias or imprecision. Discuss both direction and magnitude of any potential bias | 13-15 |
| Interpretation | 20 | Give a cautious overall interpretation of results considering objectives, limitations, multiplicity of analyses, results from similar studies, and other relevant evidence | 10-15 |
| Generalisability | 21 | Discuss the generalisability (external validity) of the study results | 14-15 |
| Other information | | | |
| Funding | 22 | Give the source of funding and the role of the funders for the present study and, if applicable, for the original study on which the present article is based | 27 |

*Give information separately for exposed and unexposed groups.

**Note:** An Explanation and Elaboration article discusses each checklist item and gives methodological background and published examples of transparent reporting. The STROBE checklist is best used in conjunction with this article (freely available on the Web sites of PLoS Medicine at http://www.plosmedicine.org/, Annals of Internal Medicine at http://www.annals.org/, and Epidemiology at http://www.epidem.com/). Information on the STROBE Initiative is available at www.strobe-statement.org.
